# The effect of medical explanations from large language models on diagnostic decisions in radiology

**DOI:** 10.1101/2025.03.04.25323357

**Authors:** Philipp Spitzer, Daniel Hendriks, Jan Rudolph, Sarah Schlaeger, Jens Ricke, Niklas Kühl, Boj Friedrich Hoppe, Stefan Feuerriegel

## Abstract

Large language models (LLMs) are increasingly used by physicians for diagnostic support. A key advantage of LLMs is the ability to generate explanations that can help physicians understand the reasoning behind a diagnosis. However, the best-suited format for LLM-generated explanations remains unclear. In this large-scale study, we examined the effect of different formats for LLM explanations on clinical decision-making. For this, we conducted a randomized experiment with radiologists reviewing patient cases with radiological images (*N* = 2020 assessments). Participants received either no LLM support (control group) or were supported by one of three LLM-generated explanations: (1) a *standard output* providing the diagnosis without explanation; (2) a *differential diagnosis* comparing multiple possible diagnoses; or (3) a *chain-of-thought* explanation offering a detailed reasoning process for the diagnosis. We find that the format of explanations significantly influences diagnostic accuracy. The chain-of-thought explanations yielded the best performance, improving the diagnostic accuracy by 12.2% compared to the control condition without LLM support (*P* = 0.001). The chain-of-thought explanations are also superior to the standard output without explanation (+7.2%; *P* = 0.040) and the differential diagnosis format (+9.7%; *P* = 0.004). We further assessed the robustness of these findings across case difficulty and different physician backgrounds such as general vs. specialized radiologists. Evidently, explaining the reasoning for a diagnosis helps physicians to identify and correct potential errors in LLM predictions and thus improve overall decisions. Altogether, the results highlight the importance of how explanations in medical LLMs are generated to maximize their utility in clinical practice. By designing explanations to support the reasoning processes of physicians, LLMs can improve diagnostic performance and, ultimately, patient outcomes.

## Introduction

Large language models (LLMs), a form of artificial intelligence (AI) capable of producing human-like text, are becoming increasingly common in clinical workflows [1, 2], including tasks such as assisting with diagnosis [3–7]. A particular promise of LLMs is to improve patient outcomes by reducing diagnostic errors [8], a major source of preventable harm [9–11], but diagnostic errors still remain a major concern, with nearly 800,000 U.S. patients dying or suffering permanent disability each year as a result [9].

Existing research has extensively evaluated the diagnostic accuracy of LLMs in clinical practice, often finding that LLMs perform as well as human physicians and, in some cases, achieve superhuman performance [5, 6, 12–14]. Other studies have examined the diagnostic accuracy of physicians when supported by LLMs compared to when they work without LLM assistance [4, 7, 12, 15]. However, these studies primarily assess the correctness of LLM-generated diagnoses but neglect how explanations provided by the LLMs influence the decision-making of physicians. Notwithstanding, a key strength of LLMs is that they can explain in natural language why a certain diagnosis seems likely. Such explanations can help physicians understand and verify the rationale behind a suggested diagnosis. Yet, the impact of different explanation formats on medical decision-making is unclear. Thus, we analyze how different formats for LLM-generated explanations influence the diagnostic accuracy of physicians.

We hypothesize that the format of LLM-generated explanations affects the diagnostic accuracy of physicians. On the one hand, explanations may improve diagnostic accuracy by helping physicians understand the reasoning behind a diagnosis. For instance, explanations with a differential diagnosis align with standard clinical practice where differential diagnoses help in reducing biases and detect rare diseases [16, 17]. Similarly, LLMs can generate explanations through chain-of-thought reasoning [18], where the LLM is asked to provide a step-by-step explanation so that a physician can verify its plausibility against their domain knowledge. On the other hand, explanations can also mislead physicians [19–21]. A key risk is automation bias, where humans overly trust machine-generated outputs such as LLM-generated advice [22, 23]. This risk is particularly pronounced for LLM-generated explanations, as LLMs tend to produce explanations that sound convincing but contain errors, a phenomenon known as “hallucinations”. Consequently, explanations, especially if detailed but incorrect, may also lead to diagnostic errors. Overall, LLM-generated explanations may support or hinder medical decision-making. However, the best-suited design of LLM-generated explanations is unclear.

Here, we analyzed the effect of different formats of LLM-generated explanations on the diagnostic accuracy of physicians (Figure 1). To do so, we conducted a randomized experiment in which radiologists were tasked to diagnose patients based on clinical vignettes containing both text and radiological images (*N* = 2020 assessments). Using a between-subject design, we varied the *explanation format*. We used the following three treatments: (1) a *standard output* that offered a diagnosis without explanation; (2) a *differential diagnosis*, which outputted the five most likely differential diagnoses in descending order; and (3) a *chain-of-thought explanation*, which detailed the reasoning process behind the diagnosis. The explanations were generated from the patient cases, involving the radiological images, using a state-of-the-art, multi-modal LLM (GPT-4; see Methods). Additionally, a control group performed the task without LLM support. We examined how the explanation formats influenced the diagnostic accuracy of physicians. Furthermore, we assessed how often doctors followed LLM advice or overrode (in)correct diagnostic suggestions. The results show that chain-of-thought explanations led to the best overall diagnostic accuracy. We further analyzed the heterogeneity in the effect across physicians and patient cases. Based on our experiment, we provide evidence-based recommendations for *how* to design explanations in medical LLMs to maximize clinical utility.

**Figure 1:**
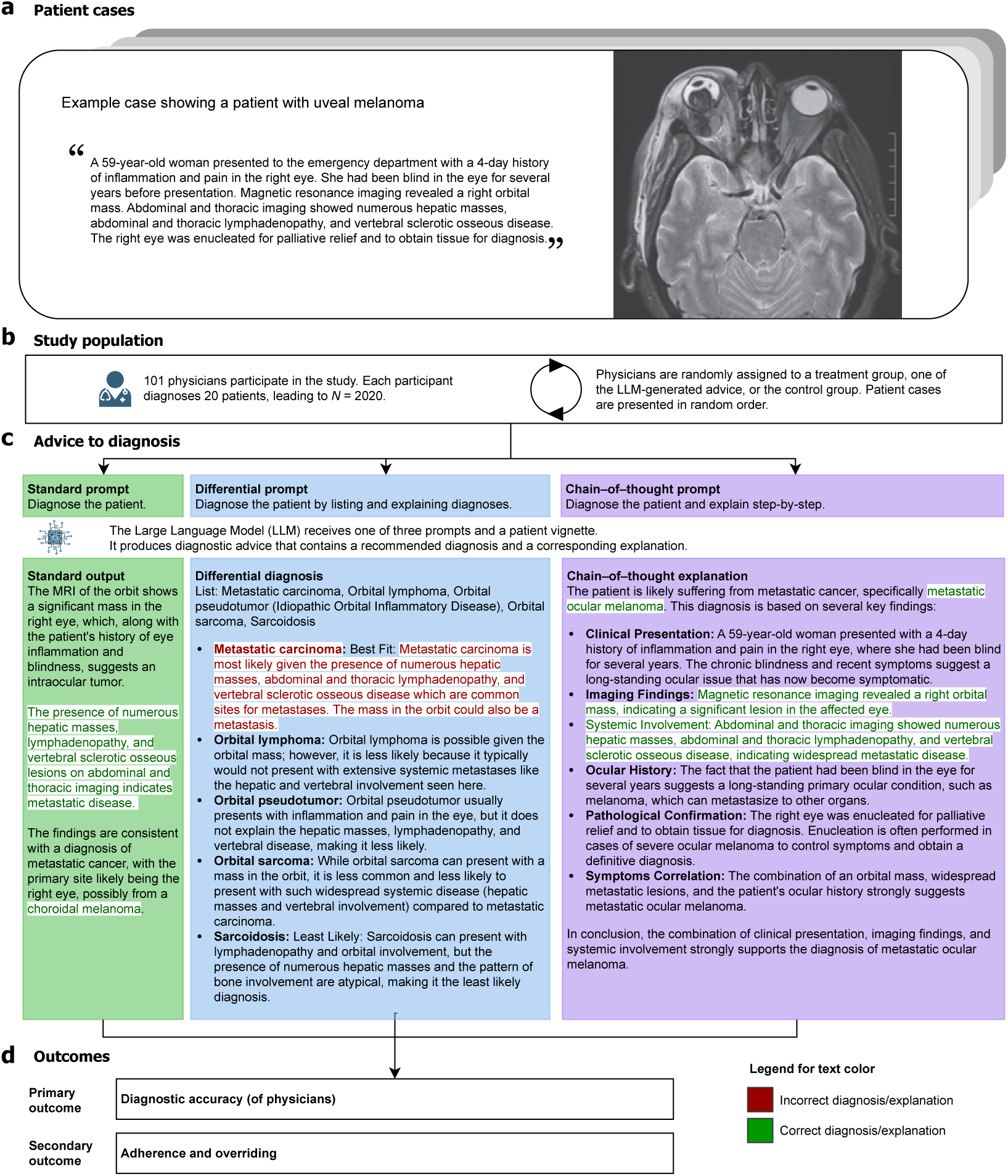
Research design. **a**, We selected 20 real-world patient cases from the *New England Journal of Medicine Image Challenge* [24]. Each patient case includes a textual description and at least one radiological image (i.e., computed tomography [CT] or magnetic resonance imaging [MRI]). **b**, We recruited 101 radiologists from the U.S. who were asked to provide diagnoses for each case and who were randomly assigned to either a control group (no LLM support) or one of three treatment groups (with LLM support). **c**, We designed prompts to produce LLM-based explanations with different formats (i.e., *standard output*, *differential diagnosis*, and *chain-of-thought*). The explanations were generated via a state-of-the-art LLM able to handle multi-modal inputs (GPT-4). However, because the LLM can make errors, both the suggested diagnoses and explanations may sometimes be incorrect. **d**, Our primary outcome is diagnostic accuracy, measured by the proportion of correct diagnoses per radiologist. To reflect clinical practice, diagnoses were collected as free-text responses (which is unlike the original *New England Journal of Medicine Image Challenge* using a multiple-choice format). As secondary outcomes, we assess how frequently radiologists adhere to correct LLM advice or overrode incorrect LLM advice.

## Results

### Experiment

To analyze how different formats of LLM-generated explanations affect diagnostic accuracy, we conducted a randomized experiment with 101 radiologists from the U.S. The radiologists had a mean of 13.6 years (standard deviation [SD] = 8.0) of medical experience. Further, the radiologists had different specializations, which we collected using a multi-option response format (e.g., general radiology: *n* = 57, abdominal imaging: *n* = 24, neuroradiology: *n* = 20, head and neck radiology: *n* = 9, cardiothoracic imaging: *n* = 8, and pediatric radiology: *n* = 5; see Supplementary Table S5)

Each radiologist was presented with 20 real-world patient cases (mean length: 55.4 words; SD = 18.8), each consisting of a brief clinical description and at least one radiological image (i.e., computed tomography [CT] or magnetic resonance imaging [MRI]), and was then asked to provide a diagnosis for the patient (*N* = 2020 assessments overall). Unlike the original *New England Journal of Medicine Image Challenge*, which uses a multiple-choice format, diagnoses were collected as free-text responses to better reflect clinical practice. For the same reason, patient cases were selected to cover a broad and diverse range of diagnostic challenges commonly encountered in radiology, following a principled inclusion process (see Methods). As a result, the cases span different radiological subspecializations (80% of patient cases can be answered using knowledge found in a standard textbook on diagnostic radiology [25], while the remaining 20% require more specialized knowledge; see Supplementary Table S6). Of note, we deliberately selected both radiologists and patient cases from different subspecializations to later assess the diagnostic accuracy in general radiology as well as across subspecializations.

Participants were randomly assigned to either a control group with no LLM support or one of three treatment groups, each with a different form of LLM advice. In the three treatment groups, physicians received advice through different types of explanations generated by GPT-4, a state-of-the-art multi-modal LLM [26, 27]. (1) *Standard output*. Here, a single best-guess diagnosis was provided, but the explanation is either absent, very brief, or paraphrasing the case description. (2) *Differential diagnosis*. Here, multiple plausible diagnoses were listed, each with a short justification, ordered by decreasing likelihood. This explanation format mirrors typical clinical reasoning, where multiple conditions are considered [28]. For the differential diagnosis format, we used two criteria for measuring diagnostic accuracy: top-1 diagnostic accuracy (the diagnosis is considered correct if the correct answer appears as the first item in the list) and top-5 diagnostic accuracy (the diagnosis is considered correct if it appears anywhere in the list of five). (3) *Chain-of-thought*. Inspired by best practice in LLM research [18], an explanation was produced that provides a step-by-step rationale of how the LLM arrives at the final recommendation, which can give physicians a deeper understanding of the reasoning process and help them identify potential errors. All outputs were kept as generated, including any diagnostic errors or hallucinations, to mimic real-world LLM use in medical practice. Example outputs for each treatment group are shown in Figure 1 (further example outputs are shown in Supplementary Figures S1 to S3). The length of the LLM-generated outputs exhibited substantial variation across the different output formats (standard output: mean = 62.7, SD = 12.5; differential diagnosis: mean = 208.6, SD = 20.2; chain-of-thought: mean = 188.6, SD = 26.2; see Supplementary Figure S5).

To quantify the quality of LLM-generated advance, we first evaluated the baseline diagnostic accuracy of the LLM output across all 20 patient cases. The LLM achieved moderate diagnostic accuracy overall(standard output: 75%; differential diagnosis: 65% for the top-1 answer and 80% for the top-5 answers; chain-of-thought: 80%; Supplementary Figure S4), which underscores the need for careful evaluation of LLM outputs and which further implies that the selected patient cases were challenging for the model. To assess the benefits of different prompting strategies for generating correct LLM outputs, we estimated a logistic regression with conditions as independent variables. The LLM with chain-of-thought prompting performed better than the standard output (coef = *−*0.288, 95% CI = [*−*1.779, 1.204], *P* = 0.705), and the LLM with chain-of-thought output performed similarly to the LLM with differential diagnosis output (coef = 0.000, 95% CI = [*−*1.549, 1.549], *P* = 1.000) (see Supplementary Table S7).

### Effect on diagnostic accuracy

Physicians augmented with LLM advice outperformed those in the control group without LLM advice (Fig. 2 for statistical comparisons based on one-sided Welch’s *t*-tests). To calculate effect sizes, we used an ordinary least squares (OLS) regression (Supplementary Table S8). Here, the diagnostic accuracy of physicians with standard output was comparable to the diagnostic accuracy of the control group (difference = 5.0 percentage points, 95% CI = [*−*1.8; 11.8], *P* = 0.150). Similarly, the diagnostic accuracy of physicians supported by a differential diagnosis was comparable to the control group (difference = 2.5 percentage points, 95% CI = [*−*4.0, 9.0], *P* = 0.446).

**Figure 2:**
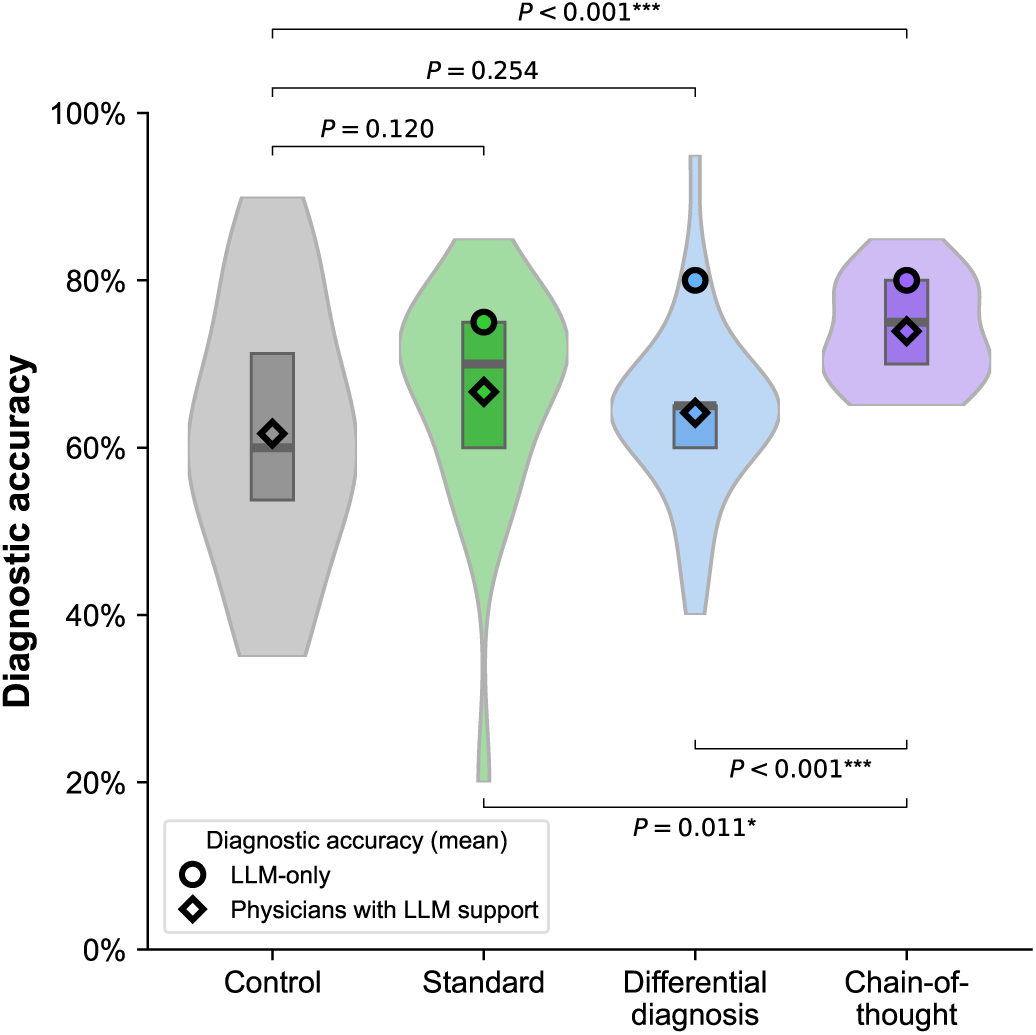
Diagnostic accuracy across different LLM explanations. The figure shows the distribution of diagnostic accuracy for all participants by condition as violin plots (which illustrate the probability density of the data). The boxplots within each violin indicate the 25% and 75% quartiles, with the median represented by the thick center line. For the differential diagnosis condition, the top-5 diagnostic accuracy is reported (the top-1 diagnostic accuracy is lower and corresponds to 65%). Statistical significance was assessed using one-sided Welch’s *t*-test to compare diagnostic accuracy between the treatment groups (in the main text, we report results from a regression analysis to obtain effect sizes).

The format of the LLM advice has a large effect on diagnostic accuracy (Fig. 2 for statistical comparisons and Supplementary Table S8 for effect sizes). Physicians in the chain-of-thought group performed best with a 12.2 percentage point improvement over the control group (95% CI = [5.3, 19.2], *P* = 0.001). Separate comparisons for the chain-of-thought versus standard condition and chain-of-thought versus differential diagnosis condition showed improvements in the outcome measure by 7.2 percentage points (95% CI = [0.3, 14.2], *P* = 0.040) and 9.7 percentage points (95% CI = [3.2, 16.3], *P* = 0.004), respectively, which confirms that the chain-of-thought condition performed best (Supplementary Table S9).

Even after controlling for various physician-specific control variables such as years of medical experience, radiology-specific expertise, hours per week spent on visual inspections, IT skills, and experience with medical AI, the effects remain robust (see Supplementary Table S10 for the regression results). Physicians with chain-of-thought explanations (difference = 15.1 percentage points, 95% CI = [7.0; 23.2], *P <* 0.001) outperformed the control group (difference = 7.0 percentage points, 95% CI = [*−*0.9; 14.9], *P* = 0.081). Further, physicians supported by chain-of-thought explanations tend to perform better than the standard output group but only at the 10% significance level (difference = 7.0 percentage points, 95% CI = [*−*0.9; 14.9], *P* = 0.081), while the comparison with the differential diagnosis group was not statistically significant (difference = 3.9 percentage points, 95% CI = [*−*3.9; 11.7], *P* = 0.325).

The findings also hold when accounting for the decision time for all cases, the length of the LLM outputs, and the length of the answers entered by participants (see Supplementary Table S11). Physicians in the chain-of-thought group significantly outperformed the control group (difference = 4.6 percentage points, 95% CI = [1.0; 8.1], *P* = 0.012), whereas physicians in the differential diagnosis condition performed significantly worse than the control group (difference = *−*4.8 percentage points, 95% CI = [*−*8.1; *−*1.5], *P* = 0.005). While the standard condition did not differ significantly from the control group (difference = 2.1 percentage points, 95% CI = [*−*3.5; 7.6], *P* = 0.467), physicians in the chain-of-thought condition also outperformed those in the standard condition (difference = 4.6 *−* 2.1 = 2.5 percentage points). Further, a breakdown of the diagnostic accuracy by patient cases is in Supplementary Fig. S7, confirming that case difficulty varies as intended based on our inclusion criteria.

To account for the baseline diagnostic accuracy across different prompting strategies, we estimated a logistic regression model where control for the correctness of the LLM diagnoses and the correctness of the LLM explanations. The standard output group and the differential diagnosis group were outperformed by the chain-of-thought group (coef = 0.504, 95% CI = [0.094, 0.914], *P* = 0.016; and coef = 1.014, 95% CI = [0.646, 1.383], *P <* 0.001; respectively) (see Supplementary Table S27).

### Mechanism of adherence and overriding

We explored adherence and overriding behavior^1^ by assessing how participants followed correct or incorrect LLM output (Fig. 3). For differential diagnosis, we assessed adherence based on the top-1 recommendation (results for top-5 recommendations are consistent; see Supplementary Figure S14). When the LLM-generated diagnosis was incorrect (Fig. 3, right column), adherence was highest in the differential diagnosis group irrespective of the correctness of the explanation (adherence for incorrect diagnosis but correct explanation: 63.3%; adherence for incorrect diagnosis and incorrect explanation: 80.0%), which also exceeded that of the standard output group (25.0% and 30.6%, respectively) and the chain-of-thought group (adherence for incorrect diagnosis and incorrect explanation: 30.4%). Note that, in the chain-of-thought group, there are no cases with an incorrect diagnosis accompanied by a correct explanation, which is expected, given the structure of the prompting strategy [18], with the aim to generate the explanation as a rationale leading to the final diagnosis. Thus, physicians in the differential diagnosis group appeared more inclined to follow the LLM-generated advice even when it was wrong.

**Figure 3:**
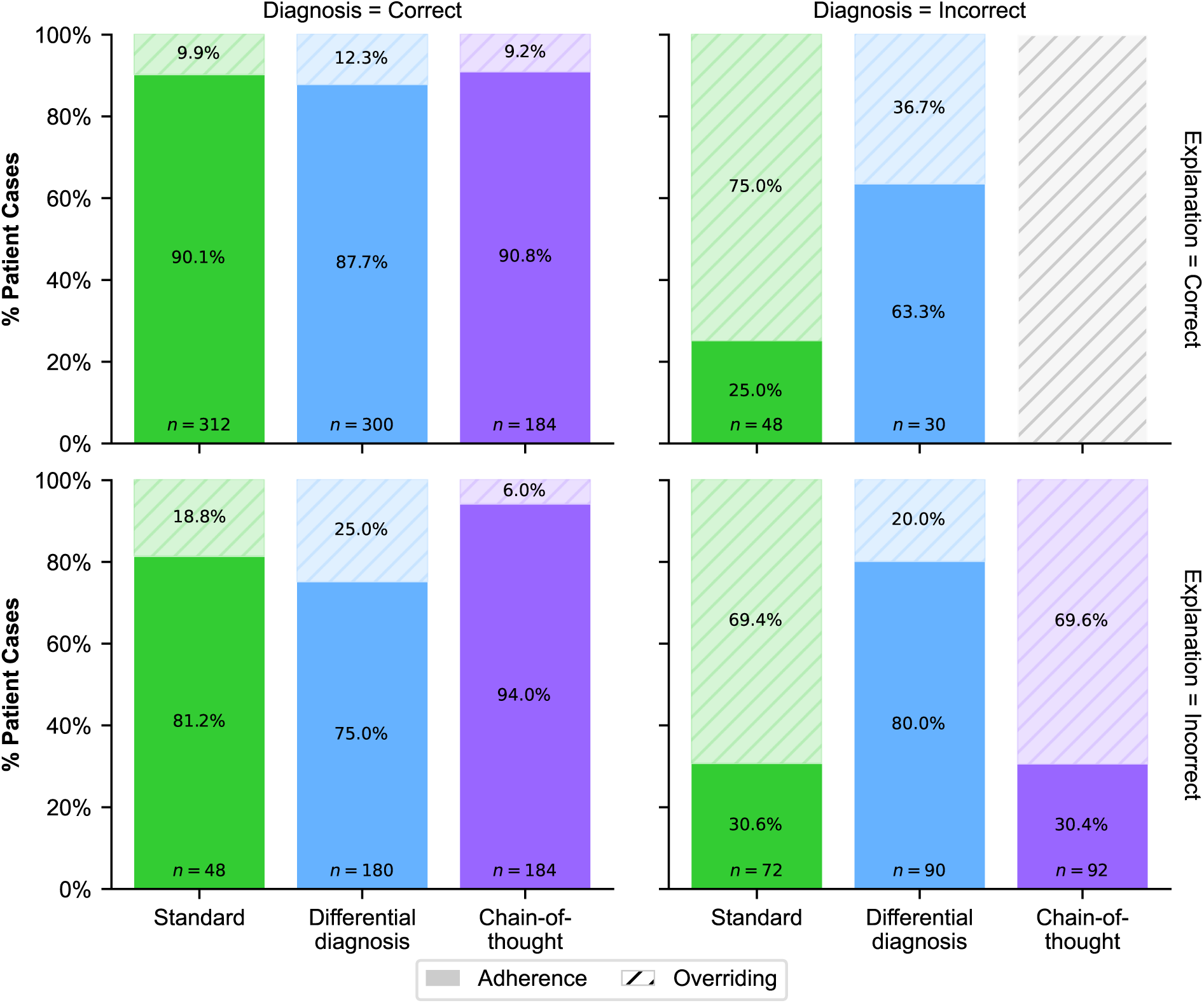
Adherence vs. overriding LLM advice. Breakdown of adherence for a correct/incorrect LLM diagnosis and for a correct/incorrect LLM explanation. Here, *adherence* was calculated as the proportion of patient cases in which participants’ diagnoses matched the LLM-generated diagnoses. Correspondingly, *overriding* was defined as 1 *− adherence*. Reported is the adherence to the top-1 answer in the differential diagnosis explanation. Results for the top-5 answers are in Supplementary Fig. S14. Specifically, the figure illustrates physician adherence to LLM-generated output, conditioned on the correctness of the diagnosis (columns) and the explanation (rows). Each panel represents a specific condition of diagnosis and explanation correctness. Comparison of the left column (diagnosis = correct) with the right column (diagnosis = incorrect) reveals that diagnostic correctness influences adherence, particularly revealing over-adherence to differential diagnoses when the recommended diagnosis is incorrect. In contrast, comparing the top row (explanation = correct) with the bottom row (explanation = incorrect) suggests that the correctness of the explanation has a limited impact on adherence across different advice types. These observations indicate that hallucinations in the diagnosis are a stronger driver of potentially inappropriate adherence than hallucinations in the explanation. Note that the condition of correct explanation and incorrect diagnosis is not represented due to a lack of generated data for this scenario, which should be expected given that chain-of-though prompting is designed to generate an explanation that matches a given diagnosis.

In contrast, when the LLM-generated diagnosis was correct (Fig. 3, left column), adherence was highest in the chain-of-thought explanations (adherence for correct explanation: 90.8%; for incorrect explanation: 94.0%), leading to more appropriate reliance on the LLM-generated advice. Here, adherence was higher compared to both the standard output group (90.1% and 81.2%) and the differential diagnosis group (87.7% and 75.0%). In sum, physicians supported with differential diagnosis explanations had consistently high adherence, even when the LLM was incorrect and thus often failed to override when the LLM advice was wrong. Conversely, the chain-of-thought explanation (in comparison to the differential diagnosis) led to more selective adherence, with physicians being more likely to follow correct LLM advice and to override incorrect LLM advice.

### Heterogeneity across physicians and patient cases

To examine heterogeneity across physicians, we analyzed diagnostic accuracy across physician subgroups with different IT skills and medical tenure (Fig. 4a,b). Overall, the chain-of-thought group tends to achieve a higher diagnostic accuracy than both the control and the differential diagnosis groups. A similar trend was observed when accounting for the complexity of patient cases (Fig. 4c).

**Figure 4:**
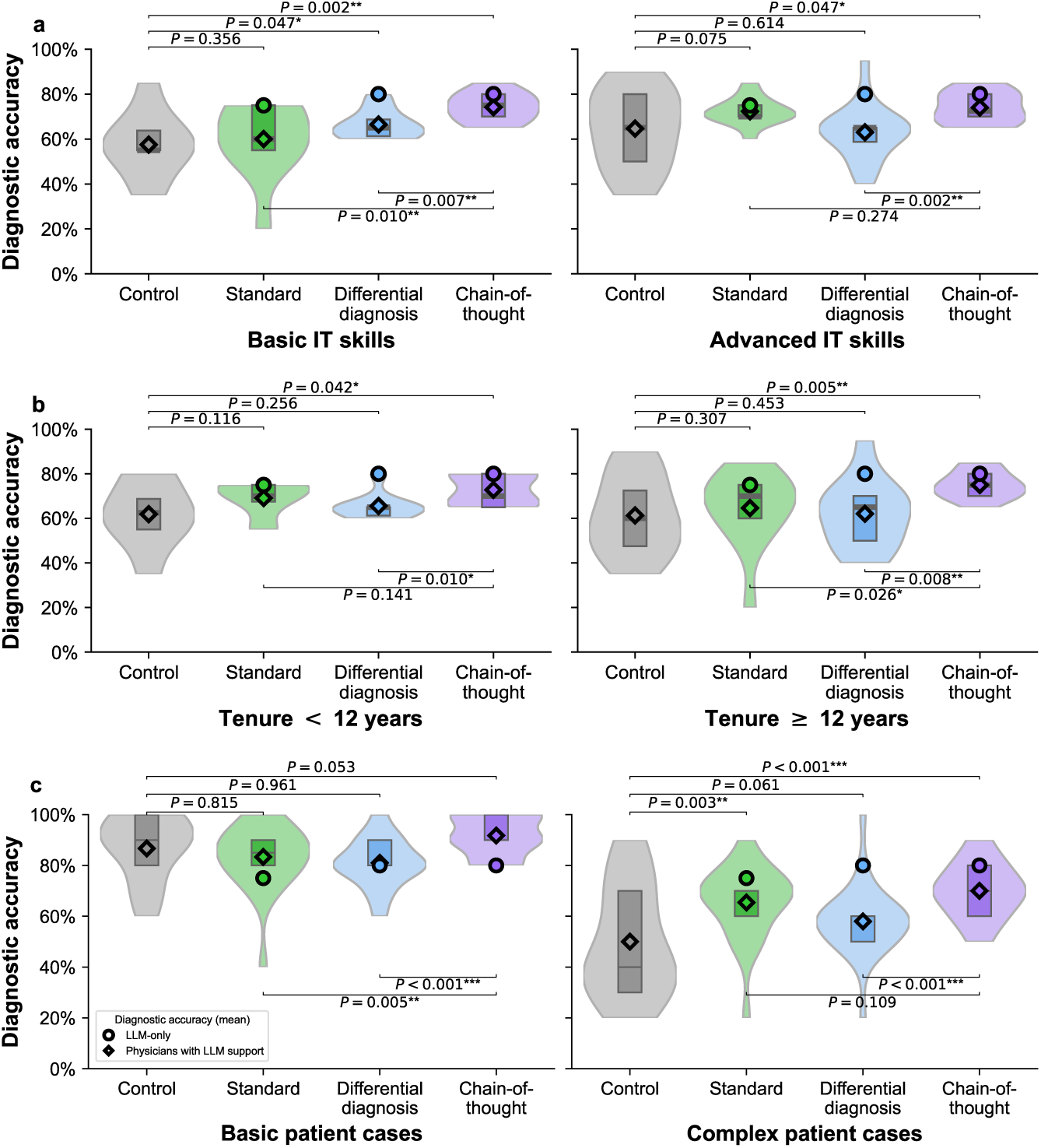
Heterogeneity across physicians and patient cases. **a**, Diagnostic accuracy was compared between different levels of IT skills, as defined by a post-hoc survey. Participants were classified as having advanced IT skills (*n* = 56) when reporting “very good”, while all others were classified as having basic IT skills (*n* = 43). **b**, Diagnostic accuracy was compared across different numbers of years with medical expertise in radiology (*n* = 46 have a tenure of *<* 12 years, while *n* = 53 have tenure of *≥* 12 years). **c**, Diagnostic accuracy for basic and complex patient cases was assessed by dividing cases into two equally sized subsets based on the median diagnostic accuracy observed in the control group, resulting in two subsets with each 10 patient cases. As such, the cases are split into subgroups based on the median diagnostic accuracy of the control group and are thus formed independently of the difficulty rating from the radiologists in our panel. The boxplots within each violin indicate the 25% and 75% quartiles, with the median represented by the thick center line. Statistical significance was assessed using one-sided Welch’s *t*-test. Estimated effect sizes using regression analysis are in Supplementary Tables S13 to S17.

### Heterogeneity across general vs. specialized radiologists

To explore the effect of explanation formats across different radiologist backgrounds, we grouped participants into general radiologists and those with subspecialization (see Fig. 5a and Supplementary Table S6). Then, we compared diagnostic performance in general radiologists (*n* = 57) against the diagnostic performance in participants with subspecialization (*n* = 66; note that participants can choose more than one area of specialization), but where we selected cases that matched the corresponding expertise.

**Figure 5:**
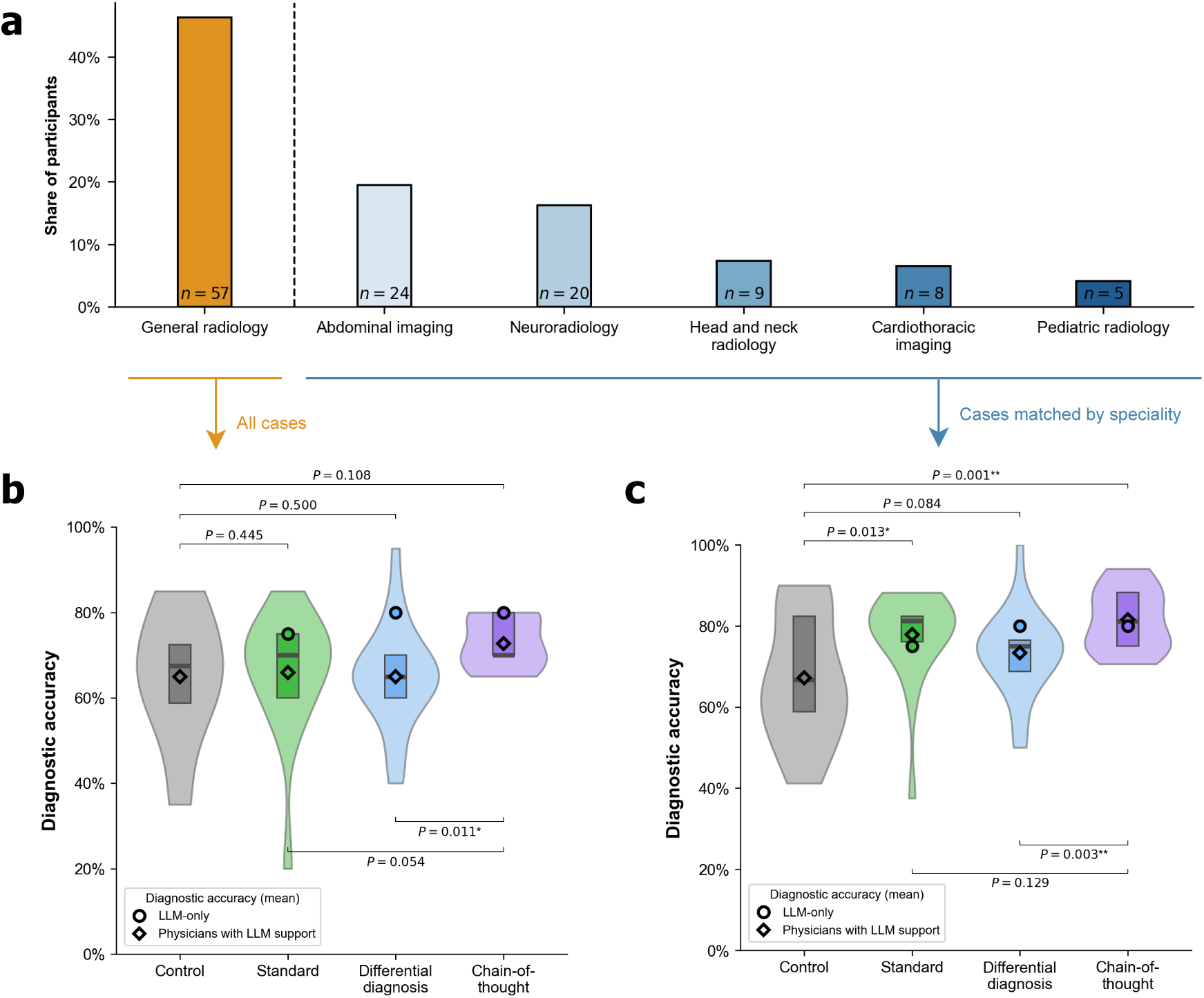
Diagnostic accuracy across different radiological backgrounds (general radiologists vs. radiologists with subspecializations). **a**, Our participant sample includes general radiologists and radiologists from different subspecializations, collected via a multi-option response format (see Supplementary Table S5). Because participants could select multiple specializations, individuals may appear in more than one subspecialty. We then matched patient cases to the radiologists based on their background, as follows: **b**, The plot shows the diagnostic accuracy assessed across all 20 patient cases for the sample of *n* = 57 general radiologists. **c**, The plot shows diagnostic accuracy for the subgroup of specialized radiologists (*n* = 66), evaluated only on patient cases matching their subspecialty. The assignment of patient cases to subspecialty is in Supplementary Table S6. The boxplots within each violin indicate the 25% and 75% quartiles, with the median represented by the thick center line. Statistical significance was assessed using one-sided Welch’s *t*-test. Estimated effect sizes using regression analysis are in Supplementary Tables S19 and S20.

While subspecialization is more common in certain regions such as the United States, many radiologists around the world continue to practice as generalists, and general radiology remains widespread in a variety of clinical settings [30–32]. We thus evaluate the diagnostic accuracy for general radiologists on all cases in our sample (Fig. 5b). Here, the diagnostic accuracy in the chain-of-thought group tends to be higher than in the standard output group but the improvement is only marginally significant (*P* = 0.0504). Interestingly, the diagnostic accuracy in the chain-of-thought group is again significantly higher than in the differential diagnosis group (*P* = 0.011) (see Supplementary Table S20 for regression results).

We next evaluate the diagnostic accuracy for specialized radiologists based on the patient cases matched by their subspecialization (Fig. 5c). While the difference between the differential diagnosis group and the standard output group was not significant (*P* = 0.129), we find that specialists in the chain-of-thought condition performed significantly better than in the control condition (*P* = 0.001) and in the differential diagnosis condition (*P* = 0.003). To understand the benefit of different explanation formats in specialized diagnostic tasks, we again performed a regression analysis (Supplementary Table S19). Specifically, we estimated the effect size of different explanation formats on the diagnostic accuracy for patient cases that matched the specific subspecialty of radiologists. Relative to the control group, improvements in diagnostic accuracy were observed for the standard output group (coef = 0.079, *P* = 0.0566) and the chain-of-thought group (coef = 0.130, *P* = 0.003) conditions. In contrast, the differential diagnosis condition showed no significant effect (coef = 0.037, *P* = 0.351). In other words, the benefit of chain-of-thought explanations over differential diagnosis explanations is not driven by radiologists’ subspecialty expertise, but is robust even for specialized radiologists.

## Discussion

The rapid adoption of LLMs in clinical settings [33] raises the question of how physicians and LLMs can most effectively collaborate to improve patient outcomes. While LLMs can help physicians verify their advice by providing explanations, the best-suited design of such explanations remains unclear. Our experiment demonstrates that the *format* of explanation plays an important role in the effectiveness of LLM support. In contrast to prior research demonstrating that LLM-augmented physicians (“human-in-the-loop”) generally outperform those without LLM support [4, 7, 12, 15, 34], we find substantial variation in the effectiveness of LLM support depending on how the information is presented. Interestingly, chain-of-thought explanations and *not* explanations with differential diagnoses provide the largest benefit to diagnostic performance. Overall, the effects were consistent across physicians with varying levels of medical experience and IT proficiency, as well as for general radiologists and those with subspecialization.

The key mechanism behind the performance gain from chain-of-thought explanations appears to be the physicians’ improved ability to assess the correctness of LLM output based on the provided reasoning. This aligns with cognitive psychology research, which suggests that structured reasoning can enhance judgment and decision-making by making thought processes more transparent [35]. The step-by-step reasoning in the chain-of-thought output helps users follow correct LLM advice (correct AI-reliance) and override flawed suggestions (correct self-reliance), consistent with research suggesting that chain-of-thought prompts help improve the reasoning of LLMs [18]. In contrast, explanations with a differential diagnosis seem to be misleading, sometimes inducing high adherence to LLM advice, even when the LLM is incorrect (often referred to as automation bias). Interestingly, prior research has already found that different diagnoses generated by LLMs may lead physicians to perform those than the LLM alone [34], yet potential solutions—such as the use of chain-of-thought explanations for clinical reasoning as proposed in our study—have not yet been explored. Notwithstanding, the differential diagnosis format may be appropriate when the LLM accuracy is high and, thus, could be more relevant in future LLMs with overall higher baseline diagnostic accuracy. Here, it would be interesting to explore how careful training of physicians may help reduce existing wrong adherence and overriding behavior. Finally, because the perceived helpfulness was similar across treatment groups (Supplementary Figure S9), the benefits from LLM advice seem unrelated to subjective factors or preferences. Instead, the benefits appear to originate from how the explanation format enables physicians to more effectively evaluate LLM advice and incorporate it into their own decision-making processes.

In general, prompt design is known to influence the correctness of LLM outputs [18]. In our study, diagnostic accuracy varied with the prompting strategy: the chain-of-thought strategy improved the accuracy of the LLM output by 5 percentage points relative to a naïve strategy with a default prompt and thus no prompt tuning. This finding aligns with best practice recommendations [18, 36] and underscores the importance of carefully fine-tuning prompts for clinical applications. Another implication of this finding is the need to boost AI literacy among physicians to help them design effective prompts, especially in light of the fact that the standard strategy for LLM deployment is one without formal training in prompt strategies [37]. Further, while the varying levels of diagnostic accuracy across different prompting strategies may have initially complicated direct comparisons, this was done intentionally and is a key strength of our study because we mimic a real-world scenario in medical LLM deployment. Nonetheless, our robustness checks confirmed that the effect of explanation formats remained robust even after adjusting for the diagnostic accuracy of the prompting strategies (see Supplementary Table S27).

Previous medical research has primarily focused on comparing the diagnostic accuracy between physicians augmented with LLM support versus those without it [4, 7, 12, 15], yet without isolating the effect of prompt design. As a result, the impact of explanation formats on diagnostic decision-making has remained unclear. Even though the importance of *how* information is presented in medical AI applications is increasingly acknowledged [38–40], empirical research focusing on decision-making is limited. Some work has explored visual annotations generated by AI tools to assist in medical image interpretation [41–43], but these studies primarily address search-and-locate tasks (e.g., lesion detection) and use outputs different from ours (e.g., heatmaps). In contrast, our study examines how different LLM explanation formats can improve diagnostic decisions.

This study has multiple limitations that present interesting directions for future research. First, we focused on only one medical specialty; however, diagnostic decisions are central to radiology and common in routine medical practice [44, 45]. Moreover, the set of patient cases covers a broad range of typical radiological assessments. Future research could explore the extent to which our findings extend to other medical specialties. Second, our analysis focused on a single time point; longitudinal studies are needed to analyze the long-term effect of LLM assistance on medical practice. Third, while we evaluated diagnostic accuracy, we did not assess the potential harm that can arise from incorrect decisions or other patient outcomes. Fourth, although some physicians in the control group reported having accessed the Internet analogous to routine practice, none indicated using LLMs, suggesting that the risk of conflated results is low. Fourth, the effects of the increased performance from chain-of-thought explanations remained statistically significant after controlling for decision time. Nevertheless, the decision time for both different diagnosis explanations and chain-of-thought explanations is longer than for the standard output group (by around 1–2 minutes), which may has economic implications (see our discussion in the Supplementary Materials). Future LLMs may generate even more informative explanations, but we also acknowledge the inherent limitations of LLMs, such as their tendency to produce hallucinations or permeating biases [46,47]. Notwithstanding, the real-world implementation of LLM in medical settings requires a careful approach considering potential risks.

Our results are based on GPT-4, a state-of-the-art multi-modal LLM for which the diagnostic accuracy closely matches that of clinical experts [6, 48, 49]. We further selected patient cases from the *New England Journal of Medicine Image Challenge* because these are expert-curated, widely recognized for their educational value, and frequently used in prior research evaluating medical LLMs [50–53]. Still, a potential limitation is the possibility that some cases may have been part of the training data for GPT-4. However, the diagnostic accuracy of GPT-4 on our task is moderate and includes frequent errors, suggesting that GPT-4 did not simply memorize the patient cases, especially as the original dataset does not include detailed explanations. Prior work also shows that small data contamination has only minimal impact on benchmark validity [54–56], and evaluating LLMs on publicly available datasets is standard practice [50,57]. Nevertheless, to avoid confounding, we refrain from drawing direct comparisons *between* LLM-supported physicians and the control group due to data overlap. However, this limitation does not affect our main findings, which rely on comparisons *within* LLM-supported conditions where explanation format—not data overlap—is responsible for observed effects.

Up to this date, misdiagnosis is a common issue in medical practice [9], including in radiology [44, 45]. While LLMs hold promise for reducing diagnostic errors [8], designing systems that effectively support collaboration with physicians remains challenging. Our findings show that the format of LLM explanations has a large impact on decision performance, thereby highlighting the need for a human-centered approach to designing real-world LLM implementations in order to maximize clinical utility and ultimately improve patient outcomes. Future LLMs may provide even more informative explanations and thereby further improve diagnostic accuracy.

## Methods

This work analyzes how the diagnostic accuracy of physicians is affected by different explanation formats of LLMs. We preregistered our hypotheses (see https://aspredicted.org/4tgb-sr3z.pdf) and tested them in a randomized experiment using a between-subject design. We complied with all local ethical regulations. The research design was approved by the Ethics Commission of LMU Munich (EK-MIS-2024-320). All participants provided informed consent.

### Procedure

To evaluate the effect of different explanation formats on the diagnostic accuracy of physicians, we designed the following experiment. First, participants received information about the study objectives. After giving informed consent, each participant was introduced to the diagnostic task and randomly assigned to one of four groups: a control group (*n* = 24) or one of three treatment groups—namely the standard output group (*n* = 24), the differential diagnosis group (*n* = 30), or the chain-of-thought group (*n* = 23). Only participants in the treatment groups received LLM-based advice. Once the participants completed the diagnostic tasks, they were asked to fill out a post-task survey. The survey collected socio-demographic information (e.g., experience in medicine in years, experience specifically in radiology in years) and asked general questions about the task (e.g., helpfulness of the LLM support). A full list of the survey items is in Supplementary Table S2.

After giving informed consent, participants had to pass a pre-registered attention check to ensure that all participants actively engaged with the task. Afterward, each participant was shown 20 cases and then asked to provide a diagnosis in the form of an open-ended text. This approach differs from the original *New England Journal of Medicine Image Challenge*, which uses a multiple-choice format. However, we opted for free-text responses to better reflect clinical practice, where diagnoses have to be generated without predefined answer options. The responses were manually coded by the author team to correct for minor typos. The cases were shown in random order to minimize bias from potential learning effects or fatigue. Each case consisted of a descriptive text plus at least one (and sometimes two) radiological images (i.e., computed tomography [CT] or magnetic resonance imaging [MRI]). Participants completed the tasks without a maximum time limit (a minimum time limit of 10 seconds was enforced for each task to ensure high-quality answers).

Our experiment was designed to reflect the diversity and heterogeneity of real-world radiology practice. To that end, we included both general radiologists and those with various subspecializations (Supplementary Table S5) and selected patient cases relevant to general radiology as well as cases spanning multiple radiological subspecialties (Supplementary Table S6). This design allowed us to assess diagnostic accuracy across different radiological backgrounds, i.e., both in general radiology and in cases where physicians engaged with content aligned with their specific subspecialty expertise.

### Patient cases

All patient cases included in this study were sourced from the *New England Journal of Medicine Image Challenge* [24], which is a high-quality collection of peer-reviewed cases, including both common and rare diagnoses, and which is used in related research to assess the diagnosis assistance from LLMs [34]. Each case consists of a short text describing the patient’s characteristics and symptoms, along with at least one medical image, such as photographs of dermatological findings, endoscopic images, histopathological slides, or radiological images. Overall, the patient cases cover both general radiology (80% of patient cases can be answered using knowledge found in core radiology educational handbooks [25]) as well as different subspecializations (20% of the patient cases require specialized expertise; see Supplementary Table S6).

In the first step, a set of only radiological cases and images were randomly sampled and the difficulty of diagnosing each case was rated by a panel of three radiologists (J.Ru., S.S., and B.F.H.) on a 5-point Likert scale. Of those, we selected 20 cases to reflect a broad spectrum of possible diagnoses and levels of difficulty, but omitted cases that were rated as either too easy or too difficult (e.g., very rare diagnoses only based on case reports). This ensured a broad and diverse set of diagnostic challenges commonly encountered in radiology. Of the 20 selected cases evaluated using a five-point Likert scale (1 = easy, 5 = difficult), 8 cases were rated as difficulty level 1, 5 cases as level 3, 5 cases as level 4, and 2 cases as level 5. Simpler patient cases included, e.g., disseminated brain metastasis or hepatic echinococcosis, while more complex cases included, e.g., disseminated mycobacterium avium complex (MAC) infection or infantile hepatic hemangiomas. The difficulty of selected cases is also reflected by the low baseline diagnostic accuracy of our control group. As such, the diversity of our patient cases is intended to mirror medical practice.

The median word count of the texts was 54.5 words, with small variability (minimum: 24 words; maximum: 85 words). Of the 20 cases, 15 include one radiological image, while the remaining 5 include two. Detailed case descriptions are provided in Supplementary Table S3 and exemplary cases are shown in Supplementary Figures S1 to S3.

### LLM advice

Participants in the treatment groups received advice generated by an LLM (GPT-4, version 2024-02-15-preview, temperature = 0.7, accessed via Microsoft Azure) based on the patient’s text and imaging data. We followed best practice in prompt design [18, 36] to produce different output formats depending on the treatment groups (see Supplementary Table S1 for the exact prompts and Supplementary Figures S1 to S3 for example outputs):

1. *Standard output* presents a single diagnosis along with either no explanation or only a concise rationale that typically mirrors the input prompt. This approach reflects LLM use cases in which no custom prompting strategy is employed. Furthermore, it aligns with the format commonly adopted in benchmarking studies from LLM research [5, 12], where the focus is primarily to assess the diagnostic accuracy of the LLM.
2. *Differential diagnosis* lists multiple possible diagnoses (here: the top five diagnoses) ordered by likelihood from most likely to less likely. This output format is motivated by that differential diagnosis is a common approach in routine medical practice [16, 17] and further acknowledges that LLMs should perform better when the top-*k* predictions are considered rather than the top-1 prediction. Intuitively, such a differential diagnosis should help physicians consider alternative diagnoses, especially for rare diseases. Unless stated otherwise, we report diagnostic accuracy for the top-5 answers, meaning that the differential diagnosis was considered correct if the correct diagnosis was contained in one of the five LLM-suggested diagnoses. Following this logic, top-1 considers the differential diagnosis correct if the *first* answer is correct. Our design choice to list *k* = 5 diagnoses in the differential diagnosis format was guided by clinical reasoning to offer a reasonable tradeoff between cognitive load for physicians and potential gains in diagnostic accuracy (see also [34] for a quantitative analysis supporting this choice). This decision is further supported by the distribution of correct answers: the correct diagnosis appeared as the top-1 suggestion in 13 out of 20 cases (65%) and within the top-2 suggestions in 16 out of 20 cases (80%), while there is only the diagnostic accuracy remained stagnant thereafter with the top-3, top-4, and top-5 suggestions containing the correct answer in 16 out of 20 cases (80% each). This pattern suggests that increasing the number of suggestions further would offer minimal incremental benefit while potentially introducing unnecessary complexity for physicians.
3. *Chain-of-thought explanation* provides a detailed, step-by-step description of the reasoning process behind how the diagnosis is made. Intuitively, this is motivated by that, on the one hand, research in machine learning suggesting that chain-of-thought prompting can improve performance [18] and, on the other hand, such format should help physicians in comparing the diagnostic process against their domain knowledge and thus identify—and eventually— override errors in the LLM advise.

We selected GPT-4 (version: 2024-02-15-preview) due to its state-of-the-art performance for diagnostic reasoning capabilities [6, 12, 13]. Further, GPT-4 is designed to handle multi-modal inputs, which allows us to process both textual descriptions and radiological images. A comparison regarding the diagnostic accuracy for another LLM is in Supplementary Figure S15, which suggests that, while other LLMs perform similarly overall, these are slightly suboptimal.

### Outcome

To evaluate the effect of different explanation formats, the primary outcome for our study was the *diagnostic accuracy* at the physician level. Specifically, we mapped the answers onto a dichotomous value (= 1 if the diagnosis is correct and 0 otherwise) and then averaged the values across all 20 patient cases for each participant. When determining whether answers were correct, we only included fully correct answers. In an additional analysis, we looked at answers, which were partially correct, but missing important details for full diagnosis (e.g., in case 2, “metabolic toxicity” is partially correct, but the underlying cause of “manganese poisoning” is missing; or, in case 3, where “gastric outlet” is partially correct, but the “superior mesenteric artery (sma) syndrome” as the cause of the obstruction is missing) and performed a robustness check where such partially correct responses were counted as correct and arrived at similar findings (see Supplementary Figure S8).

We also collected additional measures through a post-task survey to assess the perceived interactions with the LLM support (e.g., helpfulness). The survey items are listed in Table S2, which we used to analyze the perceptions of different explanations (see Supplementary Figures S9 to S13).

### Study participants

The study was conducted online (November 21, 2025 – December 1, 2025) using the online platform Qualtrics (Qualtrics International Inc.). We recruited physicians specializing in radiology from the U.S. via MSI-ACI. Participants were compensated by MSI-ACI with a fair payment above the U.S. minimum wage. No additional performance-based financial incentives were provided to preserve the ecological validity of the study and minimize potential biases due to extrinsic motivations, thereby mimicking the incentive structure of decision-making in medical practice [42].

Participants were required to meet the following inclusion criteria: They had to be licensed physicians specialized in radiology, consent to the study, and have access to a device with internet access to participate in the study. Following pre-registered exclusion criteria, we then excluded participants who did not complete the study and failed the attention check.

A total of 156 participants consented to join the study, of whom 101 completed the tasks and met the inclusion criteria, which thus forms the final sample for analysis. The reasons for exclusion were incomplete participation (*n* = 42), failure to pass the attention checks (*n* = 11), or duplicate entries (*n* = 2). The radiologists have different specializations, which we collected using a multi-option response format (e.g., general radiology: *n* = 57, abdominal imaging: *n* = 24, neuroradiology: *n* = 20, head and neck radiology: *n* = 9, cardiothoracic imaging: *n* = 8, and pediatric radiology: *n* = 5; see Supplementary Table S5). Detailed socio-demographic information and professional characteristics are reported in Supplementary Table S4.

### Statistical analysis

To compare the diagnostic accuracy across different conditions, we used analyses of variance (ANOVA) and one-sided Welch’s *t*-tests. This approach was selected due to its robustness against unequal variances and different sample sizes between groups. A significance level of *α* = 0.05 was used for all statistical comparisons.

To calculate effect sizes comparing the explanations, we estimated an OLS regression model at the physician level. The outcome was the diagnostic accuracy, modeled as a function of explanation type, with the control group as the reference and separate variables to capture the effects for groups supported by standard output, differential diagnosis, and chain-of-thought explanations. To estimate the effect of different prompting strategies on both the diagnostic accuracy of the LLM-only approach (without human involvement), we estimate a logistic regression at the assessment level. We report the coefficients on the log-odds scale for reasons of comparability.

All analyses were conducted in Python (3.11.11) using the packages `numpy` (2.2.1), `pandas` (2.2.3), `scipy` (1.15.1), `pymer4` (0.8.2), and `statsmodels` (0.14.4). The data visualizations were created with `seaborn` (0.13.2) and `matplotlib` (3.10.0).

### Robustness checks

We conducted an extensive series of robustness checks to ensure the validity and reliability of our findings. (1) We repeated the regression models while incorporating various participant-specific controls to benefit from increased power (Supplementary Table S10). (2) We further controlled for the length of participants’ answers (in the number of characters), the length of the LLM-generated advice (in the number of words), and the average duration of participants to complete the 20 patient cases (in minutes). The analysis results are shown in Supplementary Table S11. (3) To mitigate the influence of extreme outliers in our regression, we winsorized the diagnostic accuracy of study participants by limiting the extreme values in the data to the 5th and 95th percentiles (Supplementary Table S21). (4) To account for diagnostic ambiguity (e.g., when a diagnosis appears accurate but omits a specific sub-diagnosis necessary for determining the appropriate treatment strategy), our initial coding scheme classified partially correct responses as incorrect. We now repeated the analysis and counted such responses as correct (Supplementary Figure S8). (5) We estimated a mixed-effects model to control for heterogeneity across physicians and tasks (see Supplementary Table S22). (6) We estimated a quasi-binomial regression, which may limit the interpretability of the estimated effect sizes but which can better accommodate for the fact that the diagnostic accuracy is in the range between 0% and 100% (see Supplementary Table S23). (7) We controlled for the characteristics of radiological images provided in the patient cases, including the number of images in the case, the source of the image (i.e., CT or MRI), and the body region displayed in the image (i.e., abdomen, head, spine, or thorax) (see Supplementary Table S28). Across all robustness checks, the findings remained consistent with the main results.

In principle, different prompting strategies can influence the accuracy of LLM outputs. While such variability reflects real-world LLM use, potential differences in accuracy across different prompting strategies may decline with future LLMs. Thus, to remove bias from different prompting strategies, we further controlled for the diagnostic accuracy of the LLM advice within each treatment arm, which allows us to yield “ceteris paribus” estimates by factoring out variability in LLM performance (Supplementary Table S27). The approach yielded qualitatively similar conclusions.

To compare the diagnostic accuracy between general radiologists and those with subspecialization, we conducted three additional analyses: (1) We repeated the above regression analysis while controlling for the subspecialization of radiologists (see Supplementary Table S18). (2) We focused our analysis on participants who self-reported to be general radiologists and evaluated the diagnostic accuracy for this subset of participants but using all patient cases (see Supplementary Table S20). (3) We evaluated the diagnostic accuracy of specialized radioligists based on patient cases that match their expertise (see Supplementary Table S19). In all analyses, the chain-of-thought explanation resulted in the largest improvements in diagnostic accuracy.

## Data availability

De-identified data to reproduce the results is publicly available at https://github.com/ PhilippSpitzer/LLM-Explanations-for-Radiology.

## Code availability

Code to reproduce the results is available via our GitHub at https://github.com/PhilippSpitzer/ LLM-Explanations-for-Radiology. The GitHub also includes the survey of the online experiment as a Qualtrics `.qsf` file.

## Acknowledgments

The authors thank all participants.

## Author contributions

P.S. and D.H. conducted the experiment and performed the data analysis. J.Ru., S.S., and B.F.H. designed the task and assessed the correctness of participants’ responses. P.S. and D.H. coded the adherence variable. P.S., D.H., and S.F. wrote the first draft of the manuscript. All authors contributed to conceptualization, results interpretation, and manuscript editing, and approved the final manuscript.

## Supplements

### Supplementary Figures

**Figure S1:**
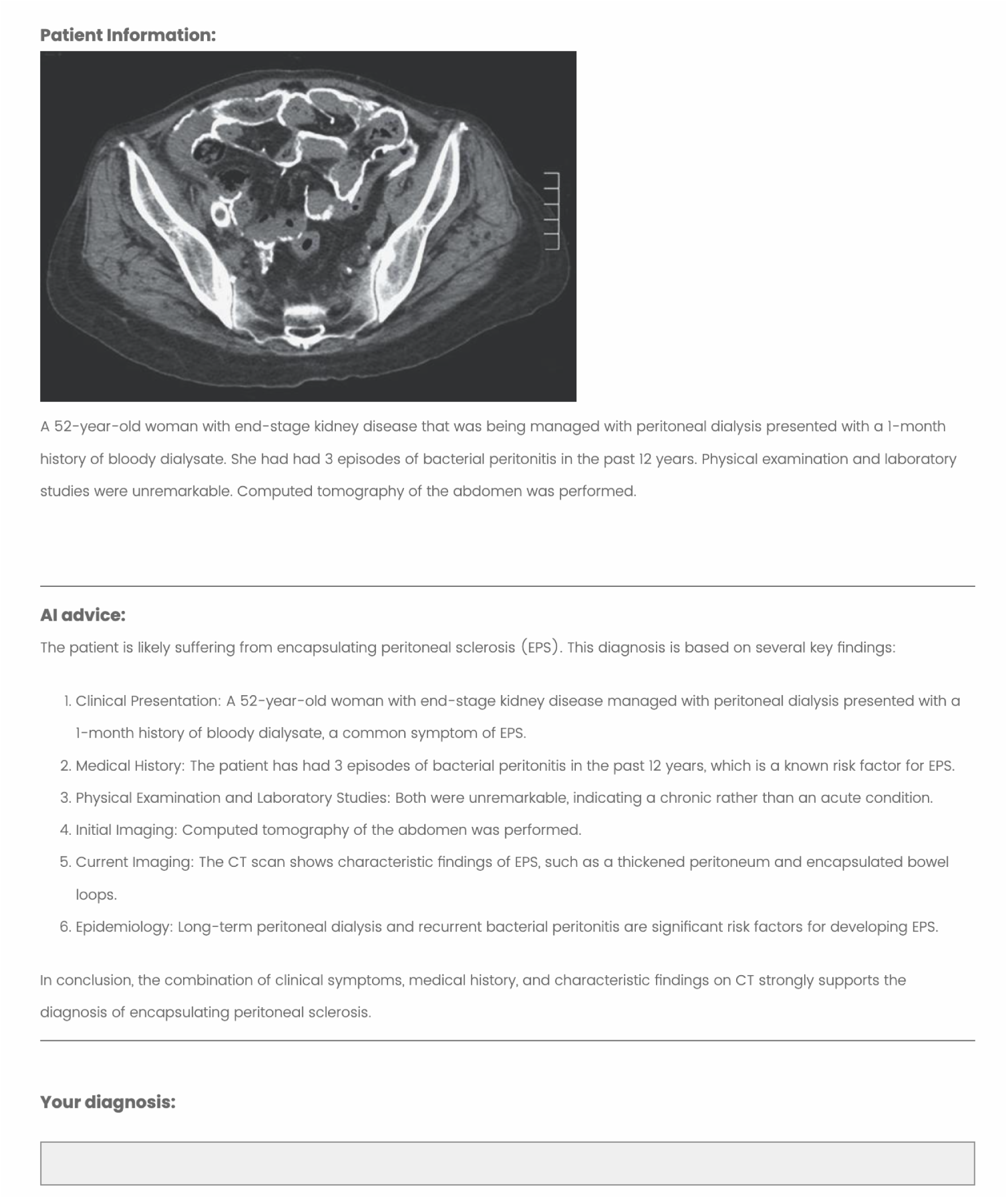
Example output: Case 5 “Encapsulating peritoneal sclerosis (EPS)”. Screenshot of the user interface during the experiment with LLM advice in the form of chain-of-thought explanations. Shown is the patient case 5, involving encapsulating peritoneal sclerosis (EPS).

**Figure S2:**
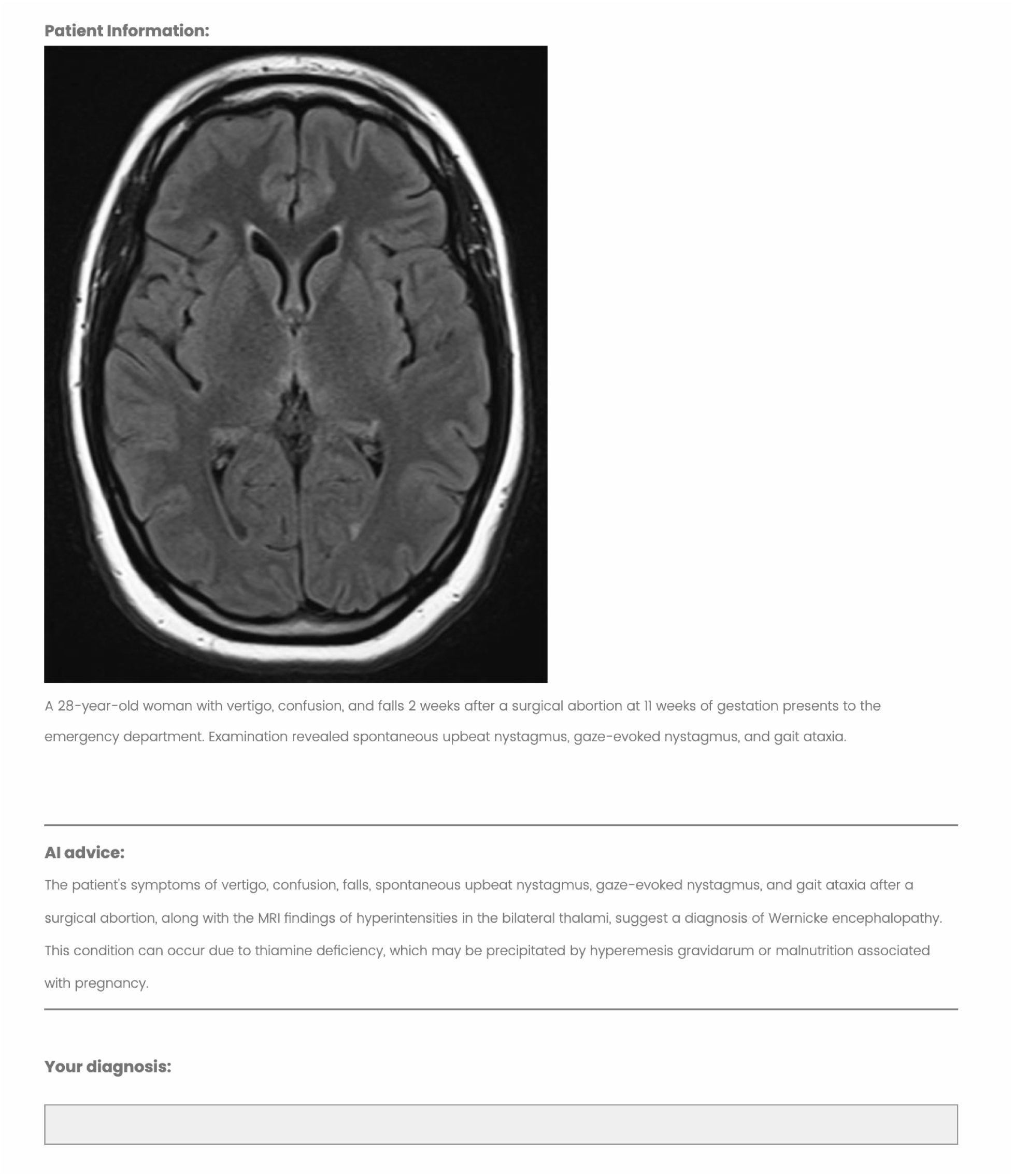
Example output: Case 14 “Wernicke’s encephalopathy”. Screenshot of the user interface during the experiment with LLM advice in the form of a standard output. Shown is patient case 14, involving Wernicke’s encephalopathy.

**Figure S3:**
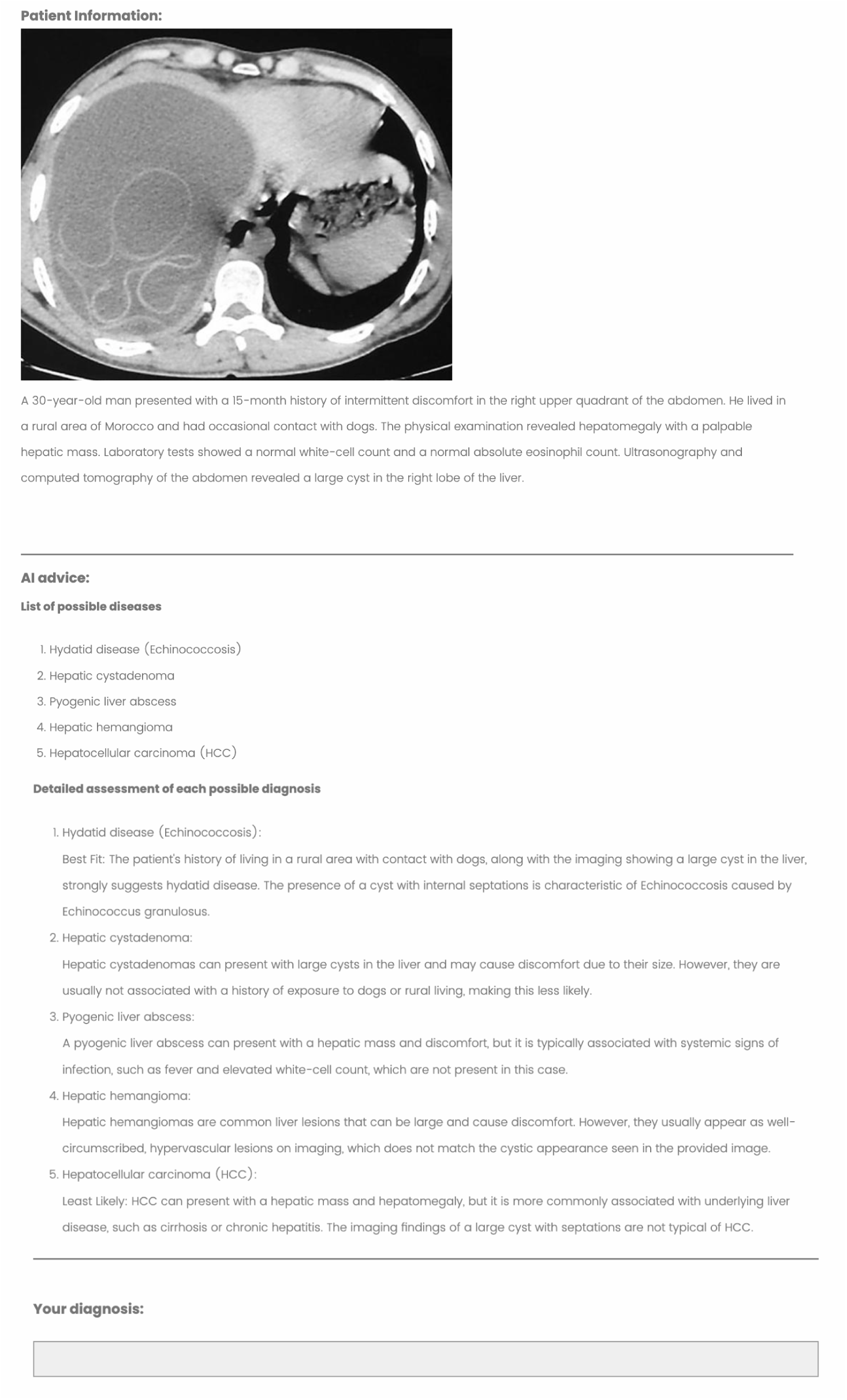
Example output: Case 18 “Cystic echinoccoccis”. Screenshot of the user interface during the experiment with LLM advice in the form of a differential diagnosis. Shown is patient case 18, involving cystic echinoccoccis.

**Figure S4:**
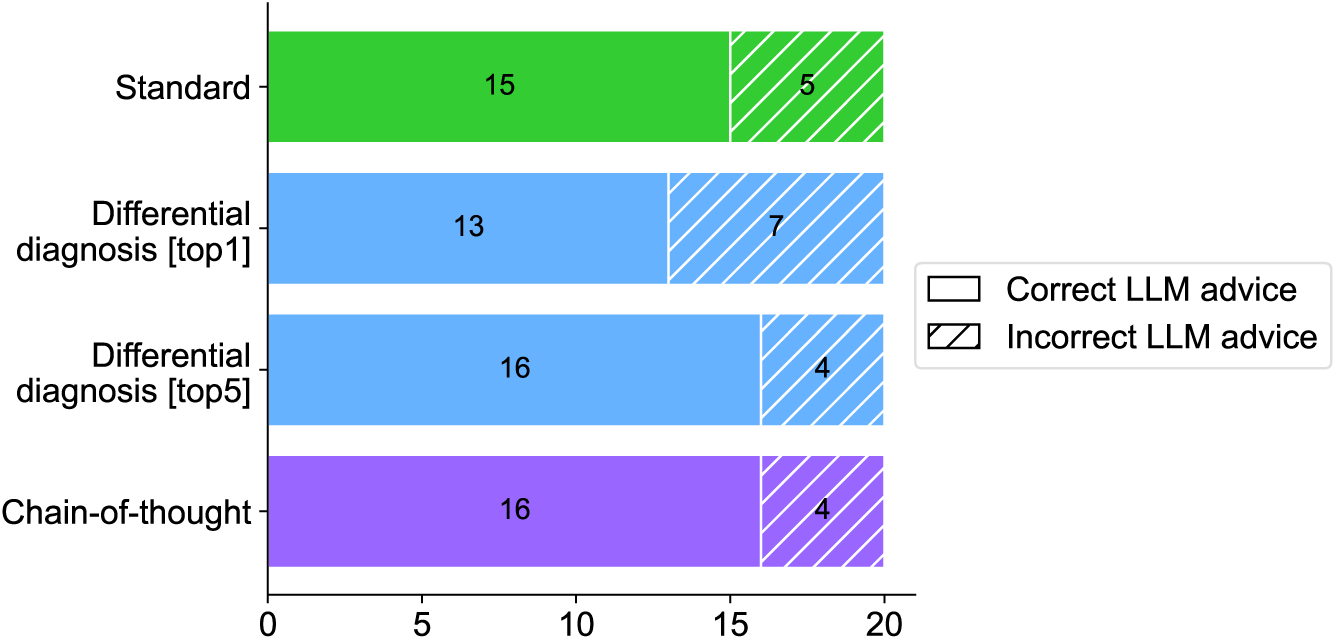
Baseline diagnostic accuracy of LLM advice. The diagnostic accuracy reports how often the LLM advice was stating the correct diagnosis (regardless of whether the explanation was correct or not). For the differential diagnosis, top-1 refers to whether the first answer was correct, while top-*k* refers to whether any of the five answers contained the correct diagnosis.

**Figure S5:**
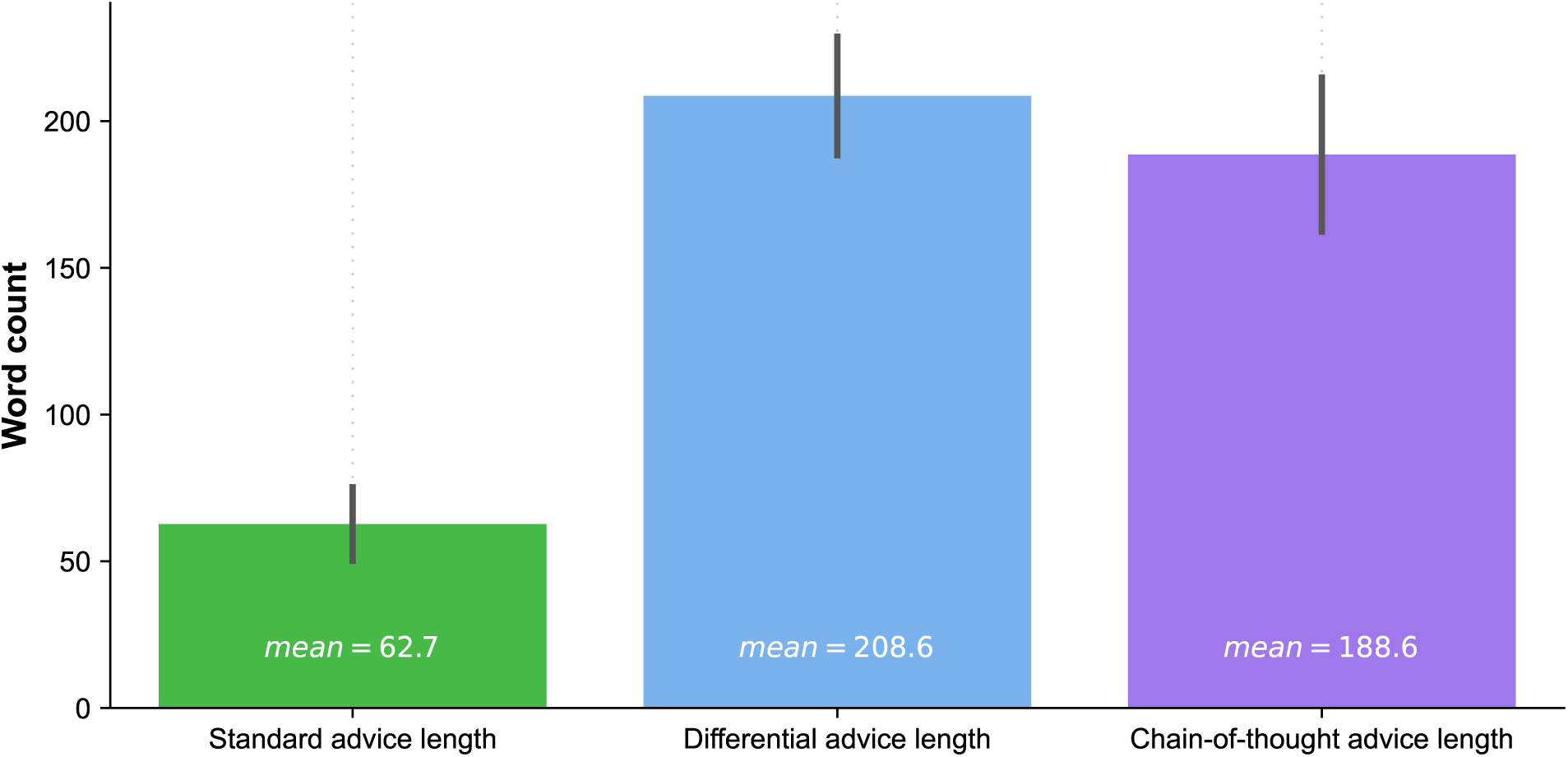
Length of LLM-generated explanations across different treatment groups (in words). Bar plots show the mean word count across all patient cases (mean as text annotations). Whiskers denote the standard deviation. For each format, we generated 20 explanations corresponding to the *n* = 20 cases.

**Figure S6:**
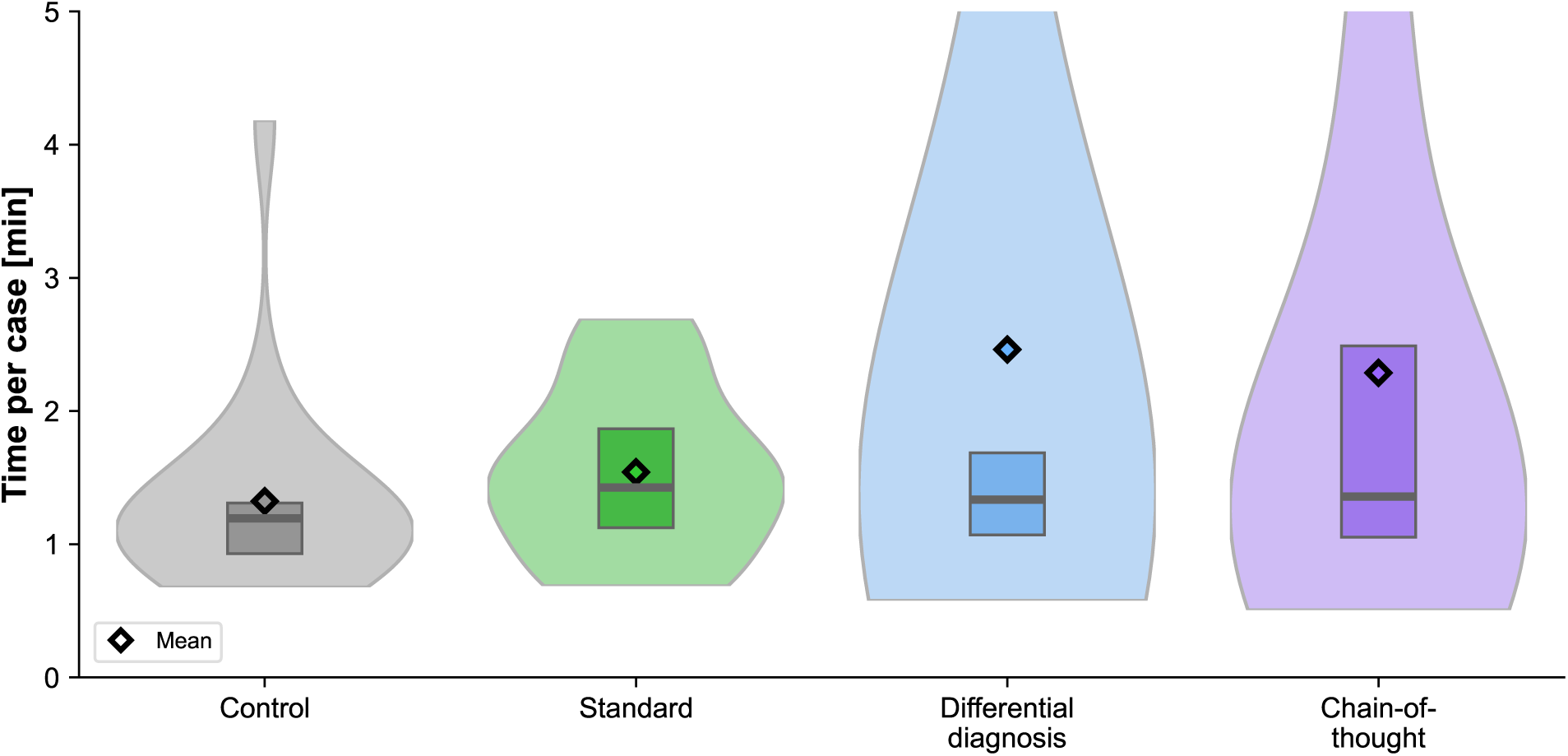
Decision time. Distribution of average time per case over all participants in minutes. The figure shows the distribution as violin plots (which illustrate the probability density of the data). The boxplots within each violin indicate the 25% and 75% quartiles, with the median represented by the thick center line and the mean by a diamond. Each condition contains *n* = 20 observations.

**Figure S7:**
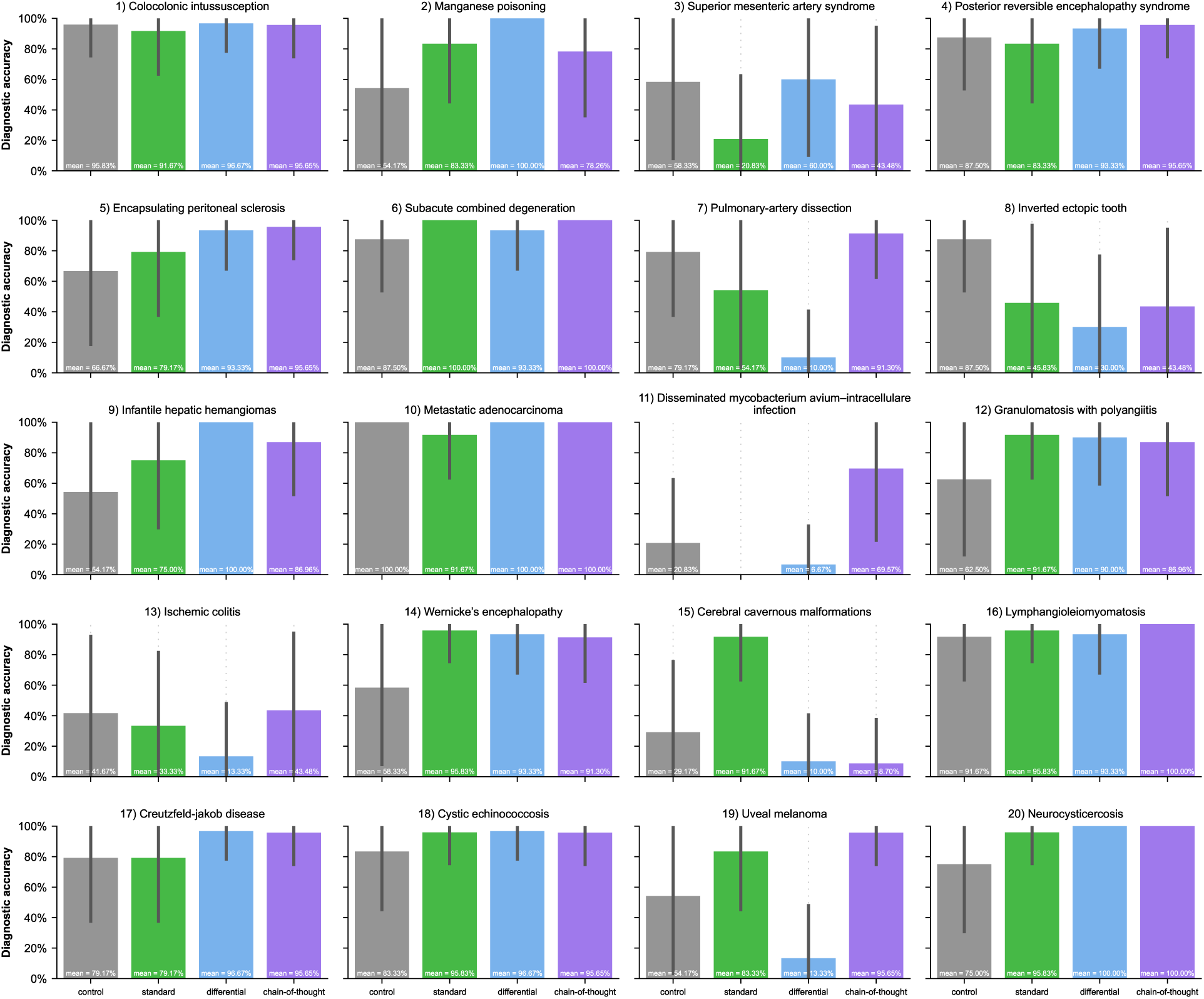
Diagnostic accuracy across patient cases. For certain cases, such as *collagenous enterocolitis* and *superior mesenteric artery syndrome*, all conditions led to high diagnostic accuracy rates above 80%. However, other patient cases proved more challenging, with notably lower performance across all conditions. For instance, for the cases of *ischemic colitis* and *cerebral cavernous malformations*, diagnostic accuracy rates dropped below 40% for several conditions. The chain-of-though group showed particularly strong performance in complex cases like *drug-induced thrombocytopenia* and *disseminated mycobacterium avium-intracellulare infection*, where other conditions often led to incorrect diagnoses. Physicians supported by the standard output performed well only in cases such as *Wernicke’s encephalopathy* and *cystic echinococcosis*, while the differential diagnostic explanation showed strengths in diagnosing *pulmonary-artery dissection* and *infantile hepatic hemangioma*. This variation in performance across different medical conditions highlights the complexity of medical diagnosis and suggests that different prompting strategies might be more effective for different types of cases. Whiskers refer to standard deviations. For each patient case, the sample size by conditions is as follows: *n* = 24 for the control group, *n* = 24 for the standard condition, *n* = 30 for the differential diagnosis conditions, and *n* = 23 for the chain-of-thought condition.

**Figure S8:**
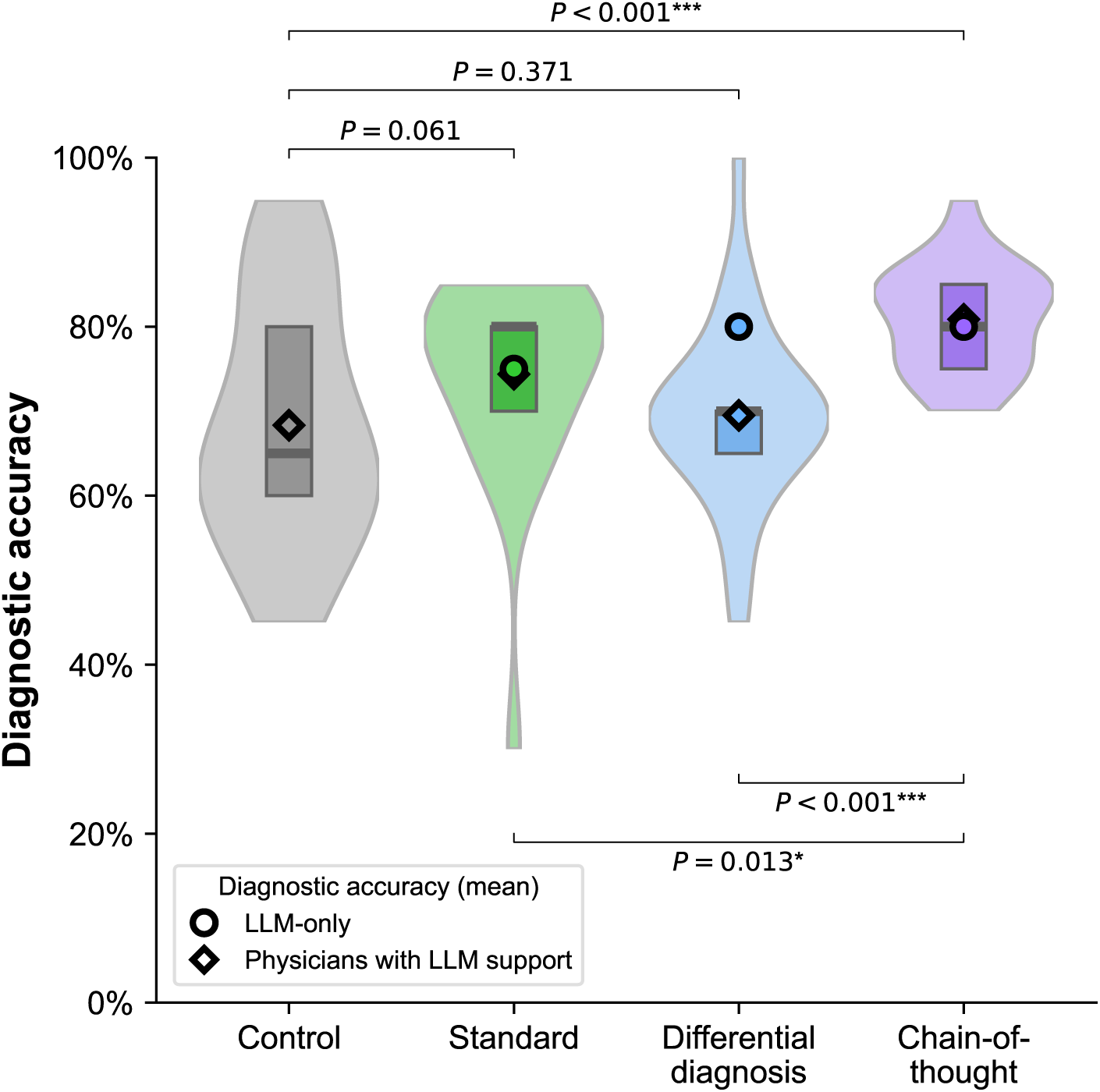
Robustness check for the diagnostic accuracy when counting partial responses as correct. To account for diagnostic ambiguity (for instance, when a diagnosis appears accurate but omits a sub-diagnosis necessary for determining an appropriate treatment strategy), we perform a robustness check. The coding scheme in the main paper classified such partial responses as incorrect, while we now present an analysis in which we classified such responses as correct. The figure shows the distribution of diagnostic accuracy for all participants as violin plots (which illustrate the probability density of the data). The boxplots within each violin indicate the 25% and 75% quartiles, with the median represented by the thick center line. For the differential diagnosis condition, the top-5 diagnostic accuracy is reported (the top-1 diagnostic accuracy is lower and numbers to 65%). Statistical significance was assessed using one-sided Welch’s *t*-test to compare diagnostic accuracy between the treatment groups. The sample size is *n* = 101 aggregated at the participant level and, hence, does not contain repeated measures (*n* = 24 for the control group, *n* = 24 for the standard condition, *n* = 30 for the differential diagnosis conditions, and *n* = 23 for the chain-of-thought condition).

**Figure S9:**
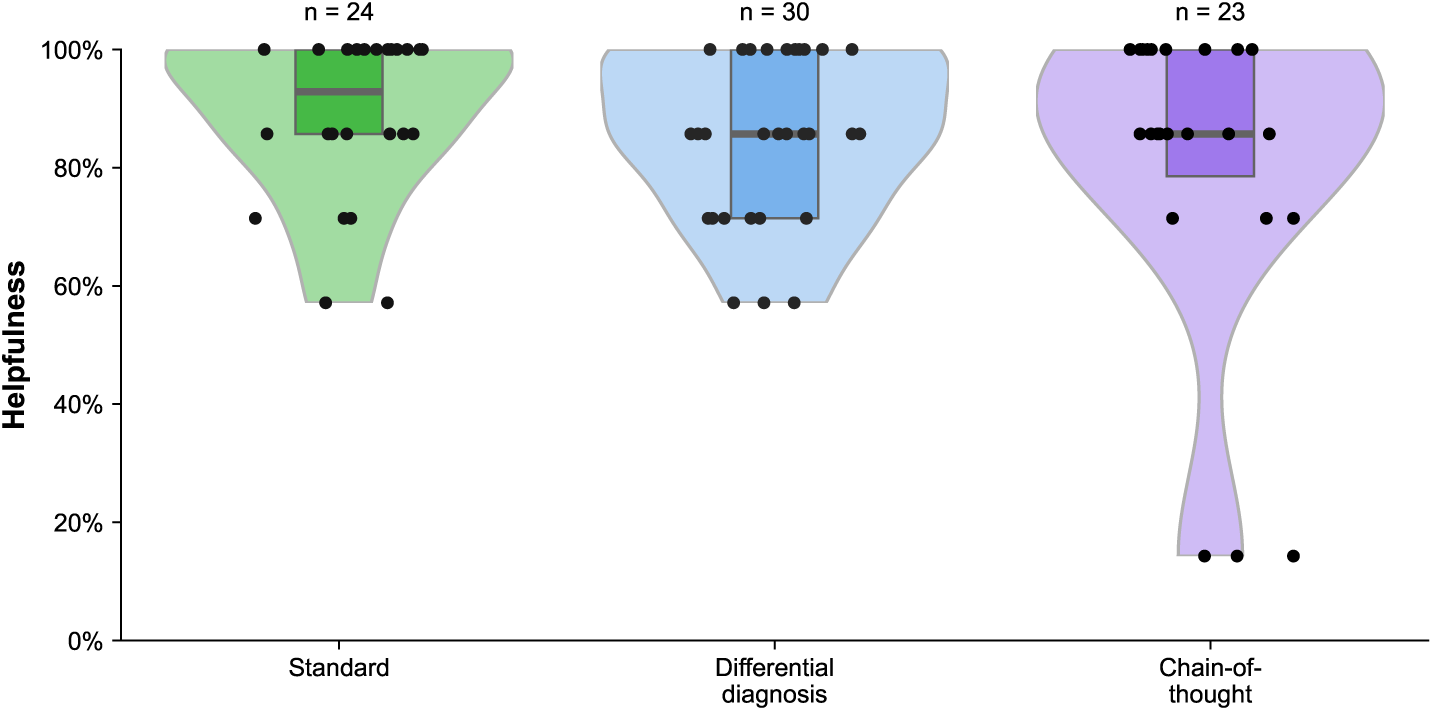
Effect on helpfulness. The survey items were combined by averaging the Likert-scale ratings per participant and normalizing the result to a percentage scale. Helpfulness was not collected for the control group, leading to *n* = 77. Helpfulness was assessed by participants after completing the diagnostic tasks. An ANOVA reveals no statistically significant differences between the conditions (*F* -value = 1.324, *P* -value = 0.271), indicating that the type of explanation had no effect on helpfulness. Statistical significance was assessed using one-sided Welch’s *t*-tests, showing no significant differences between any of the conditions (all *P* -values *>* 0.05). Whiskers denote standard deviations, there are no repeated measures.

**Figure S10:**
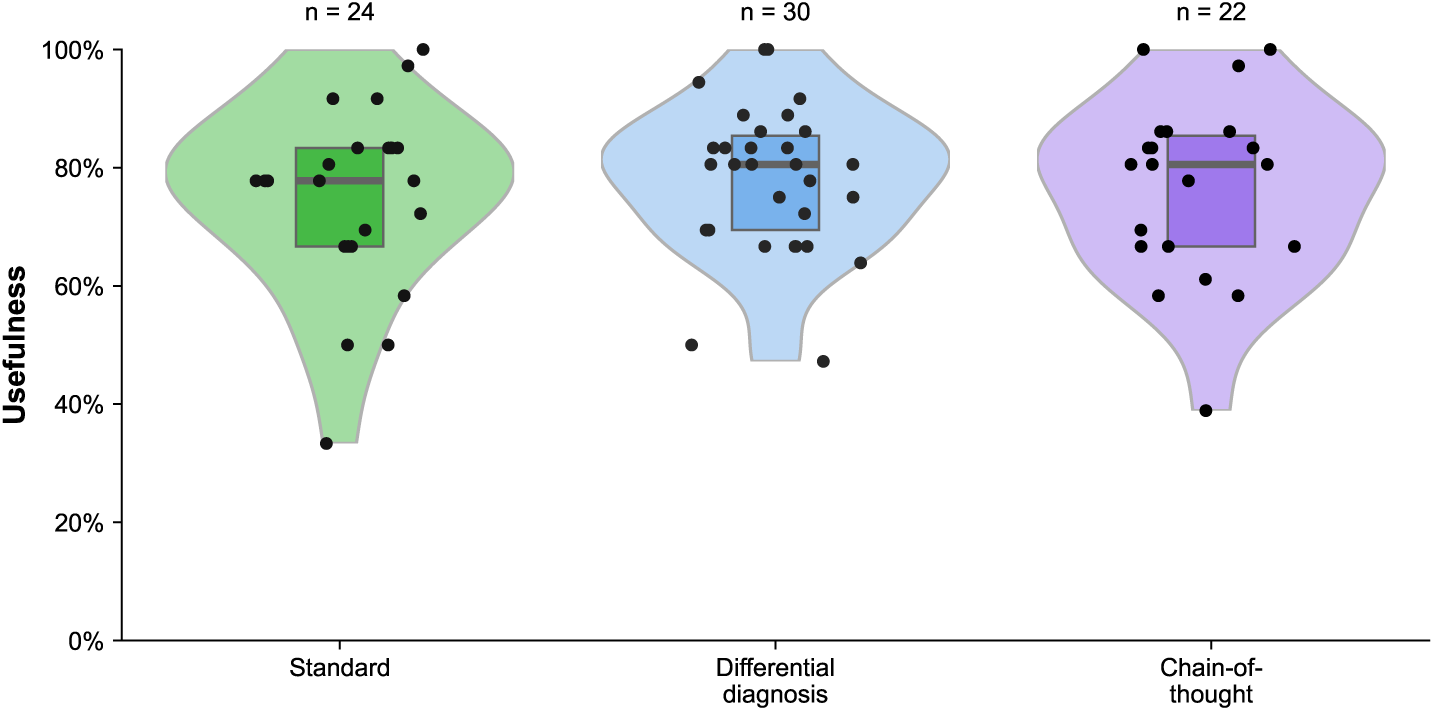
Effect on usefulness. The survey items were combined by averaging the Likert-scale ratings per participant and normalizing the result to a percentage scale. The sample size of the post-study questionnaire varies, as the questionnaire was not fully displayed in two cases where participants selected specific specialties (“neuroradiologist” and “interventional radiology”). Usefulness was not collected for the control group, leading to *n* = 76. An ANOVA analysis reveals no statistically significant differences between the conditions (*F* -value = 0.690, *P* -value = 0.505), indicating that the explanation type had no effect on usefulness. Statistical significance was assessed using one-sided Welch’s *t*-tests, showing no significant differences between any of the conditions (all *P* -values *>* 0.05). Whiskers denote standard deviations, there are no repeated measures.

**Figure S11:**
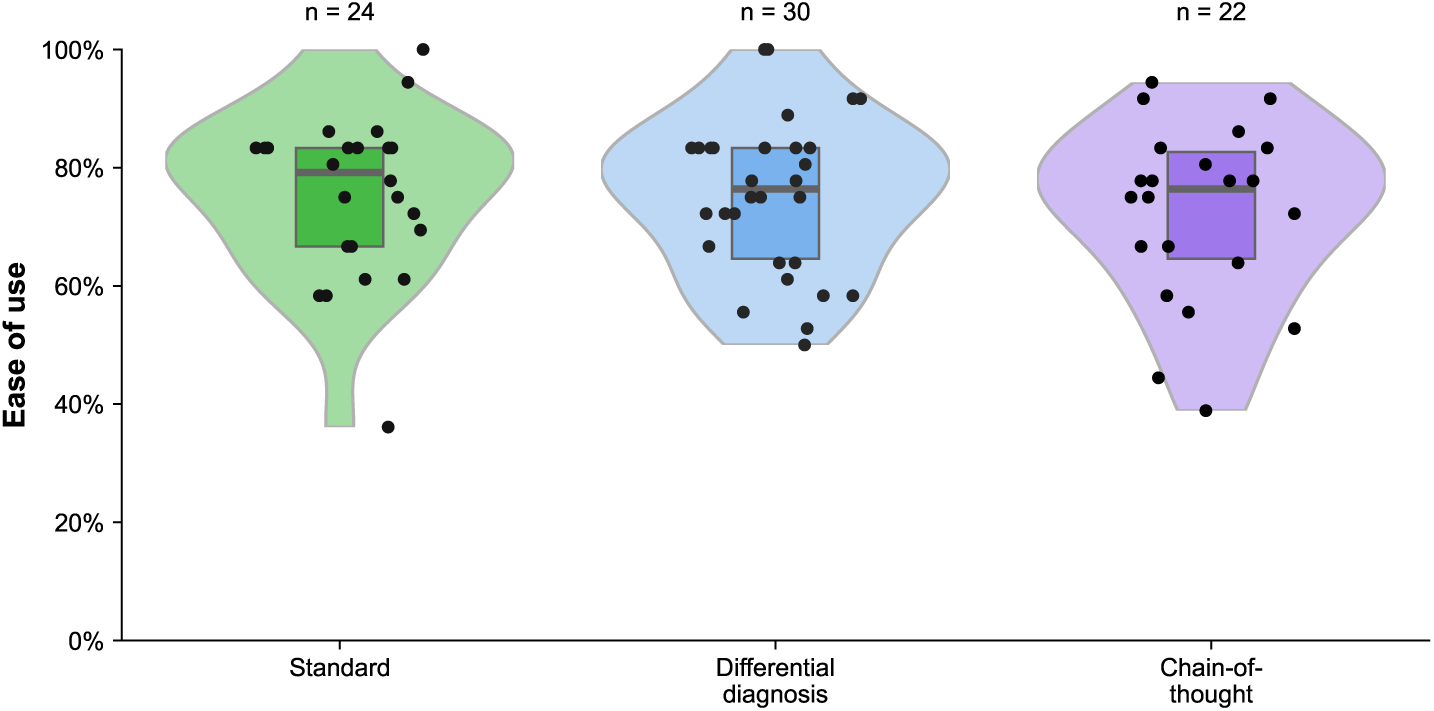
Effect on ease of use. The survey items were combined by averaging the Likert-scale ratings per participant and normalizing the result to a percentage scale. The sample size of the post-study questionnaire varies, as the questionnaire was not fully displayed in two cases where participants selected specific specialties (“neuroradiologist” and “interventional radiology”). Ease of use was not collected for the control group, leading to *n* = 76. An ANOVA analysis reveals no statistically significant differences between the conditions (*F* -value = 1.313, *P* -value = 0.275), indicating that the explanation type had no effect on ease of use. Statistical significance was assessed using one-sided Welch’s *t*-tests, showing no significant differences between any of the conditions (all *P* -values *>* 0.05). Whiskers denote standard deviations, there are no repeated measures.

**Figure S12:**
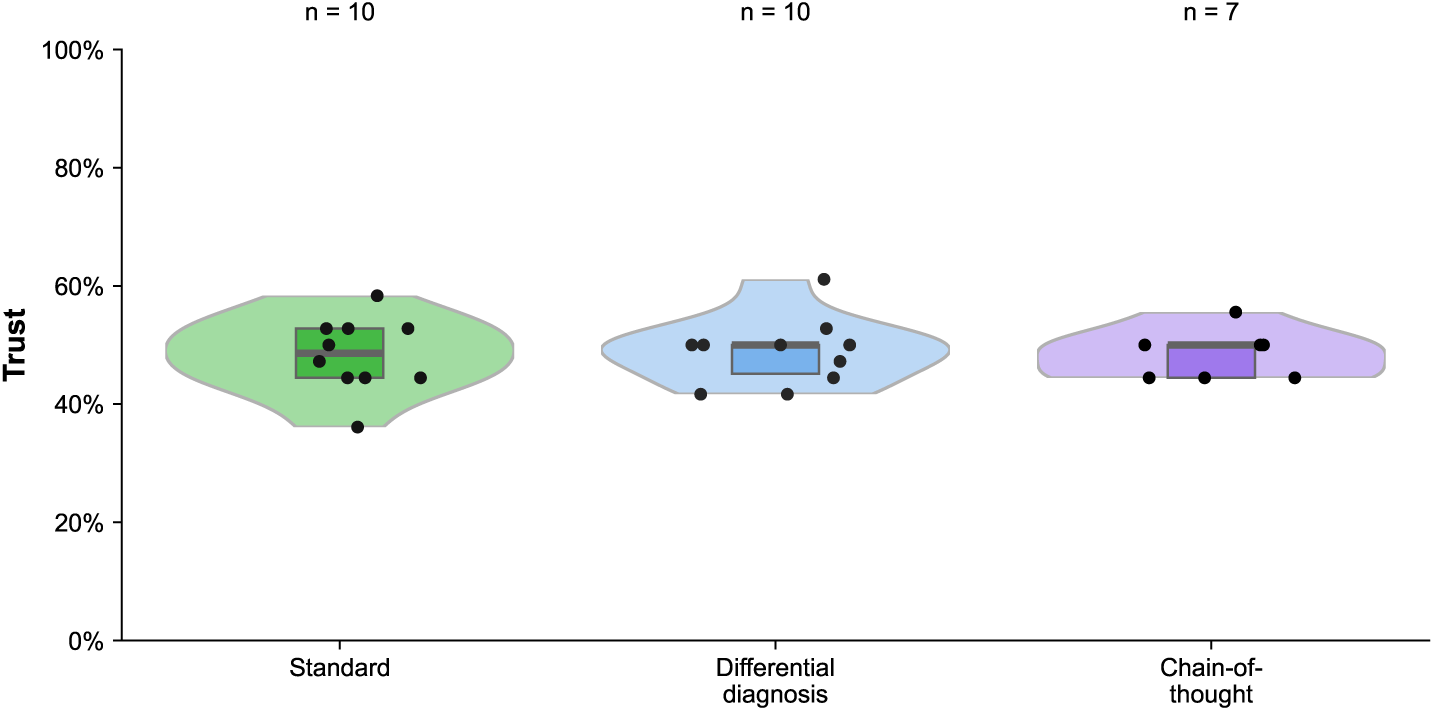
Effect on trust. The survey items were combined by averaging the Likert-scale ratings per participant and normalizing the result to a percentage scale. The sample size for the trust construct is smaller due to a technical error in the study’s setup, resulting in missing data. An ANOVA reveals no statistically significant differences between the conditions (*F* -value = 0.333, *P* -value = 0.718), indicating that the explanation type had no effect on trust. Statistical significance was assessed using one-sided Welch’s *t*-tests, showing no significant differences between any of the conditions (all *P* -values *>* 0.05). Whiskers denote standard deviations, there are no repeated measures.

**Figure S13:**
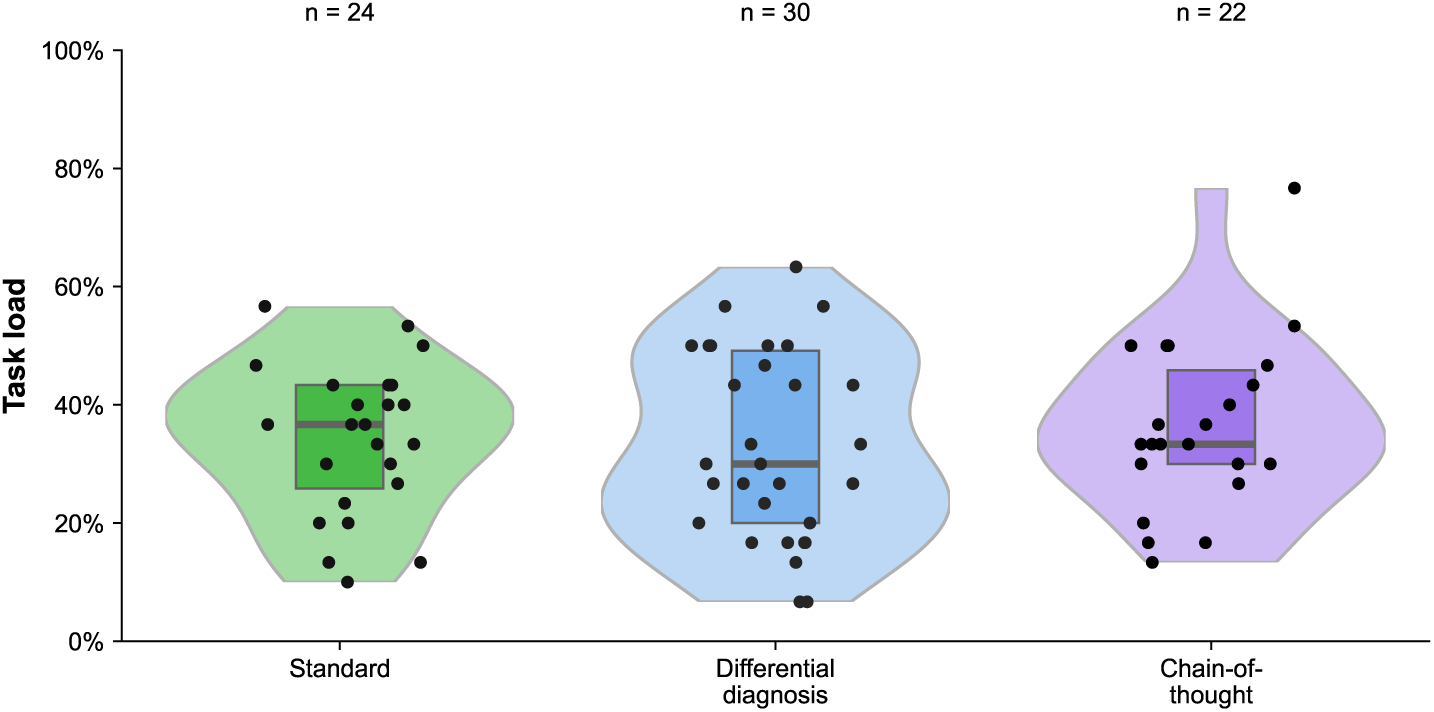
Effect on task load. The survey items were combined by averaging the Likert-scale ratings per participant and normalizing the result to a percentage scale. The sample size of the post-study questionnaire varies, as the questionnaire was not fully displayed in two cases where participants selected specific specialties (“neuroradiologist” and “interventional radiology”), leading to *n* = 99. An ANOVA reveals no statistically significant differences between the conditions (*F* -value = 0.505, *P* -value = 0.679), indicating that the type of explanation had no effect on task load. Statistical significance was assessed using one-sided Welch’s *t*-tests, showing no significant differences between any of the conditions (all *P* -values *>* 0.05). Whiskers denote standard deviations, there are no repeated measures.

**Figure S14:**
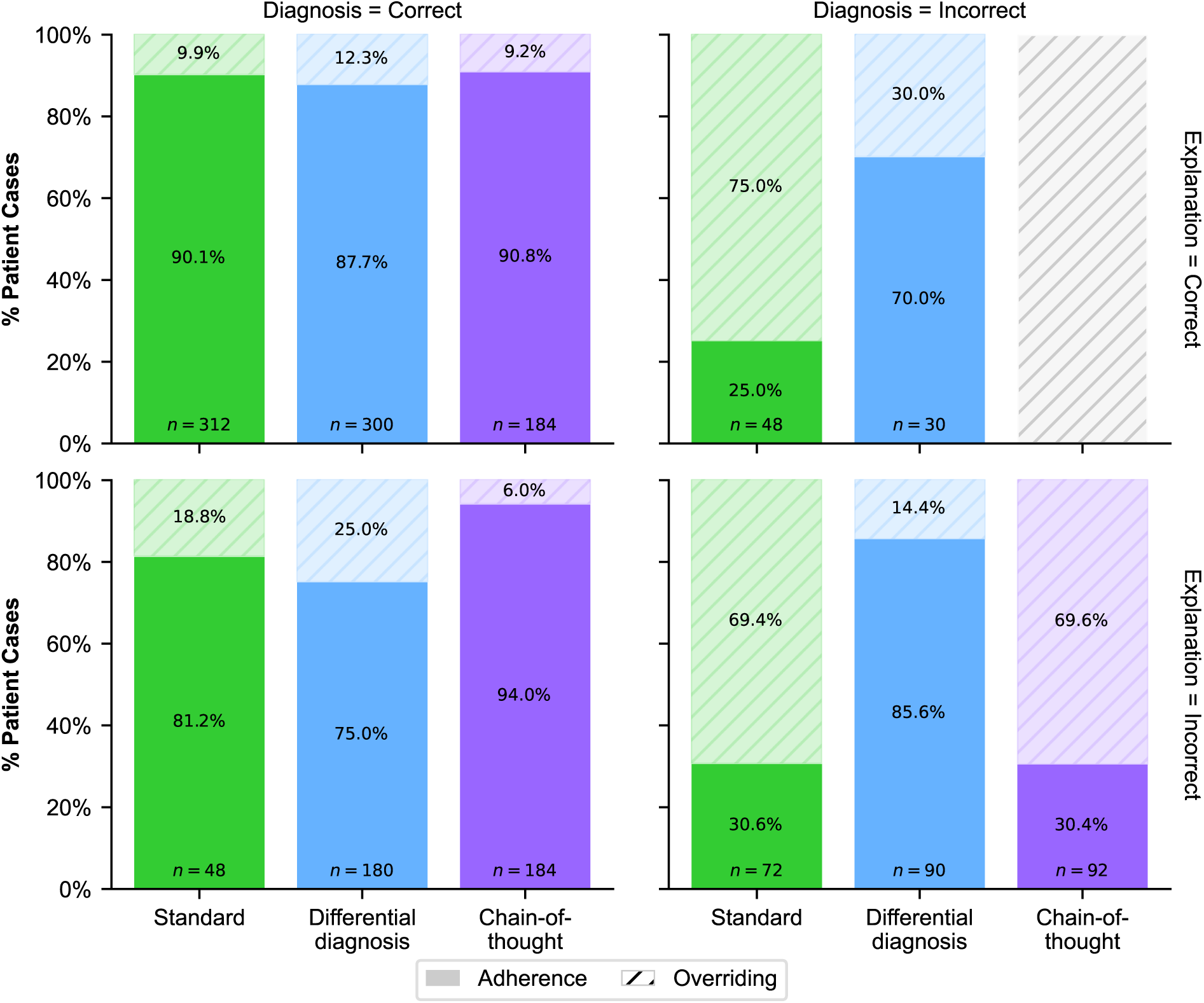
Adherence vs. overriding LLM advice for different explanation formats. The figure depicts the breakdown of physician adherence (solid fill) and overriding (pattern fill) behaviors relative to LLM-generated diagnostic recommendations. Adherence was defined as the proportion of patient cases in which physicians’ diagnoses followed the LLM-generated diagnoses, with overriding calculated as 1*−adherence*. Panels illustrate adherence by the correctness of the recommended diagnosis (columns) and correctness of the explanation provided by the LLM (rows), across the three conditions (standard, differential diagnosis, and chain-of-thought). Notably, diagnostic correctness of the LLM strongly influences adherence rates, highlighting increased over-adherence when diagnoses were incorrect, which is especially evident in the differential diagnosis condition. Conversely, the correctness of explanations shows only little impact on adherence patterns, suggesting that inaccuracies (hallucinations) in diagnoses contribute more significantly to inappropriate adherence than inaccuracies in explanations. The *correct explanation and incorrect diagnosis* scenario is absent as no LLM-generated advice met these criteria within the chain-of-thought group, which can be expected due to that chain-of-thought is designed to amend a given diagnosis with an explanation.

**Figure S15:**
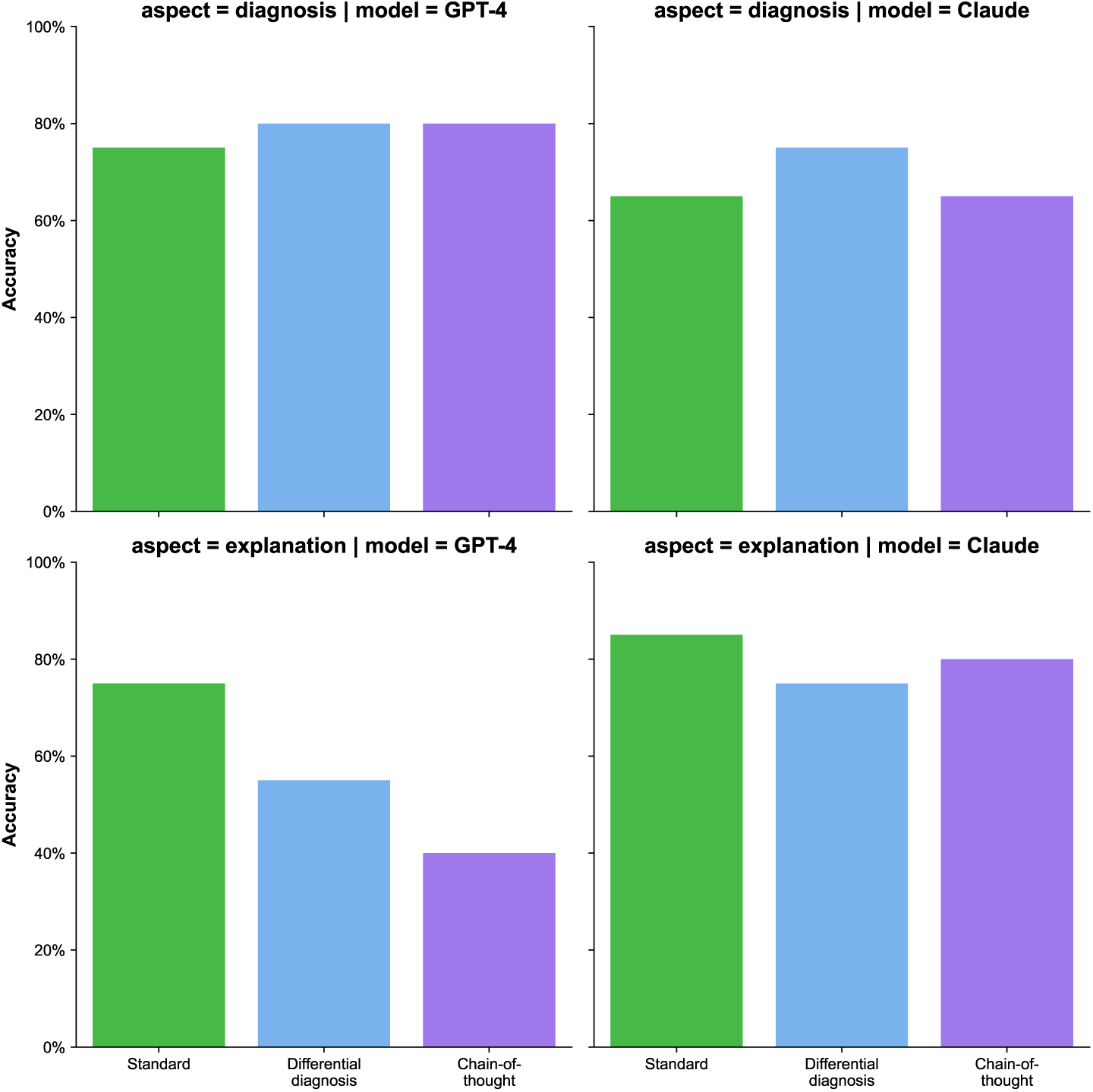
Comparison of GPT-4 and Claude in diagnosing patients. To provide a justification for our choice of GPT-4, we now compare the diagnostic accuracy of GPT-4 against another state-of-the-art LLM, namely, Claude. The reason for the choice is that Claude is designed to handle multi-modal input (i.e., patient descriptions with radiology images). This is unlike many other open-source LLMs (e.g., Llama-3, DeepSeek-R1), which, to this date, are limited to text-only input. We generated advice for the 20 patient cases from our study with images and text using Claude (version claude-3-opus-20240229, temperature = 1.0, maximum token generations = 4096). We then assessed the accuracy of both the diagnoses and the explanations provided by the model. Overall, both GPT-4 and Claude are comparable in terms of diagnostic accuracy. The largest difference is observed in the accuracy of observations. Considering that the effect of incorrect explanations on the participants in our study was small, it is reasonable to assume that we would obtain similar results from our study using a different LLM. To further support this claim, we calculated the inter-rater reliability of the models in diagnosing patient cases. The analysis of the inter-rater agreement between GPT-4 and Claude using Cohen’s kappa revealed a fair agreement on both diagnoses (*κ* = 0.243) and explanations (*κ* = 0.348). The sample size for each condition on each subplot is *n* = 20, there are no repeated measures.

### Supplementary Tables

**Table S1:**
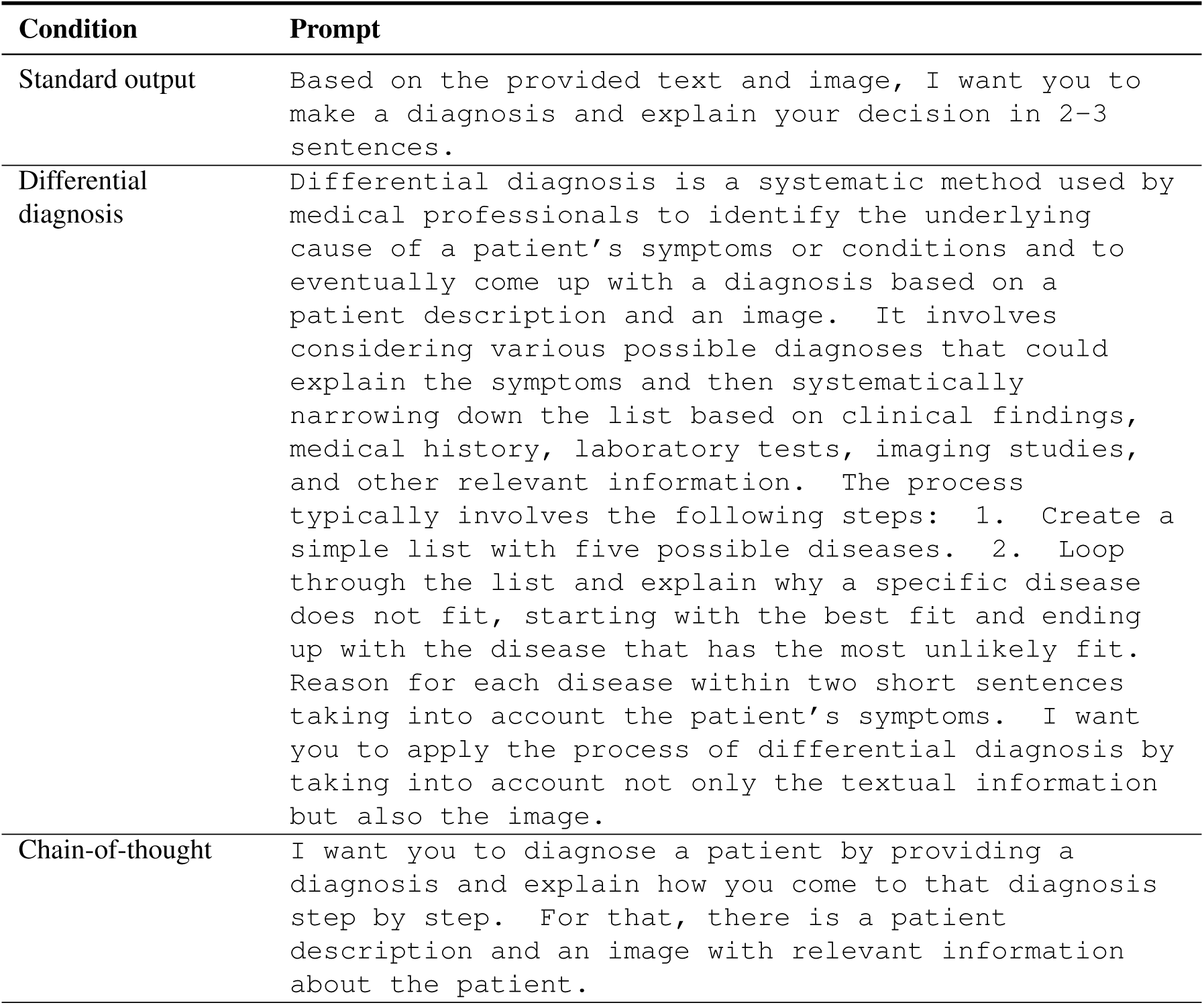
Prompts for generating explanations. Shown are the three different prompting approaches for generating medical diagnosis explanations: standard output, differential diagnosis, and chain-of-thought reasoning

| Condition | Prompt |
| --- | --- |
| Standard output | Based on the provided text and image, I want you to make a diagnosis and explain your decision in 2-3 sentences. |
| Differential diagnosis | Differential diagnosis is a systematic method used by medical professionals to identify the underlying cause of a patient's symptoms or conditions and to eventually come up with a diagnosis based on a patient description and an image. It involves considering various possible diagnoses that could explain the symptoms and then systematically narrowing down the list based on clinical findings, medical history, laboratory tests, imaging studies, and other relevant information. The process typically involves the following steps: 1. Create a simple list with five possible diseases. 2. Loop through the list and explain why a specific disease does not fit, starting with the best fit and ending up with the disease that has the most unlikely fit. Reason for each disease within two short sentences taking into account the patient's symptoms. I want you to apply the process of differential diagnosis by taking into account not only the textual information but also the image. |
| Chain-of-thought | I want you to diagnose a patient by providing a diagnosis and explain how you come to that diagnosis step by step. For that, there is a patient description and an image with relevant information about the patient. |

**Table S2:** Items in the post-task survey. Survey items measure participants’ socio-demographics, expertise, and perceptions toward the LLM advice in the medical diagnosis tasks.

| Variable | Items | Scale |
| --- | --- | --- |
| Explanation helpfulness | How helpful were the explanations?<br>How can we make the AI advice more helpful for diagnosing patients? | 7-point Likert-scale and free-form text answer respectively |
| Expertise in Generative AI | How do you rate your expertise in using Generative AI like ChatGPT? | 7-point Likert-scale |
| Specialization | What is your specialization in medicine?<br>Which area do you specialize in? | Multiple-choice-question with option for free-form text answer |
| Medical experience | How many years of experience do you have as a medical professional? | Number |
| Experience in radiology | How do you rate your experience in radiology? | 7-point Likert-scale |
| IT skills | How strong would you consider your IT skills? | 7-point Likert-scale |
| Experience with medical AI systems | How much experience do you have in working with medical AI systems? | 7-point Likert-scale |
| Weekly visual inspection time | How many hours per week do you spend on visual inspections? | 7-point Likert-scale |
| Perceived self-efficacy | Please estimate the percentage (0–100%) of diagnoses that you made correctly.<br>How successful were you in accomplishing what you were asked to do? | Free-form text answer with check for numeric inputs and 7-point Likert-scale respectively |
| Perceived AI efficacy | Please estimate the percentage (0–100%) of diagnoses that the AI system made correctly.<br>How likely is the AI system to make a bad estimate? | Free-form text answer with check for numeric inputs and 7-point Likert-scale respectively |
| AI expectations | How well did the AI system perform in comparison to your expectations? | 7-point Likert-scale |
| Usefulness | • Using the AI system would enable others to classify architectural styles of buildings more quickly.<br>• Using the AI system would improve the performance when classifying architectural styles of buildings.<br>• Using the AI system would increase productivity for classifying the architectural styles of buildings.<br>• Using the AI system would enhance my effectiveness in classifying the architectural styles of buildings.<br>• Using the AI system would make it easier to classify the architectural styles of buildings.<br>• I would find the AI system useful for classifying the architectural styles of buildings. | 7-point Likert-scale |
| Ease of use | • Learning to operate the AI would be easy for me.<br>• I would find it easy to get the AI to do what I want it to do.<br>• My interaction with the AI would be clear and understandable.<br>• I would find the AI to be flexible to interact with.<br>• It would be easy for me to become skillful at using AI.<br>• I would find the AI easy to use. | 7-point Likert-scale |
| Trust | • The AI is deceptive.<br>• I am suspicious of the AI's intent, action, or outputs.<br>• The AI's actions will have a harmful outcome.<br>• I am confident in the AI.<br>• The AI is reliable.<br>• I can trust the AI. | 7-point Likert-scale |
| Task load | • How mentally demanding was the task?<br>• How physically demanding was the task?<br>• How hurried or rushed was the pace of the task?<br>• How hard did you have to work to accomplish your level of performance?<br>• How insecure, discouraged, irritated, stressed, and annoyed were you? | 7-point Likert-scale |
| Open-ended questions | • Have you used any assistive technology/tools (internet research, books, colleagues, etc.) in filling out this survey?<br>• In case you clicked "yes", which assistive technology/tools did you use? | Free-form text answer |

**Table S3:**
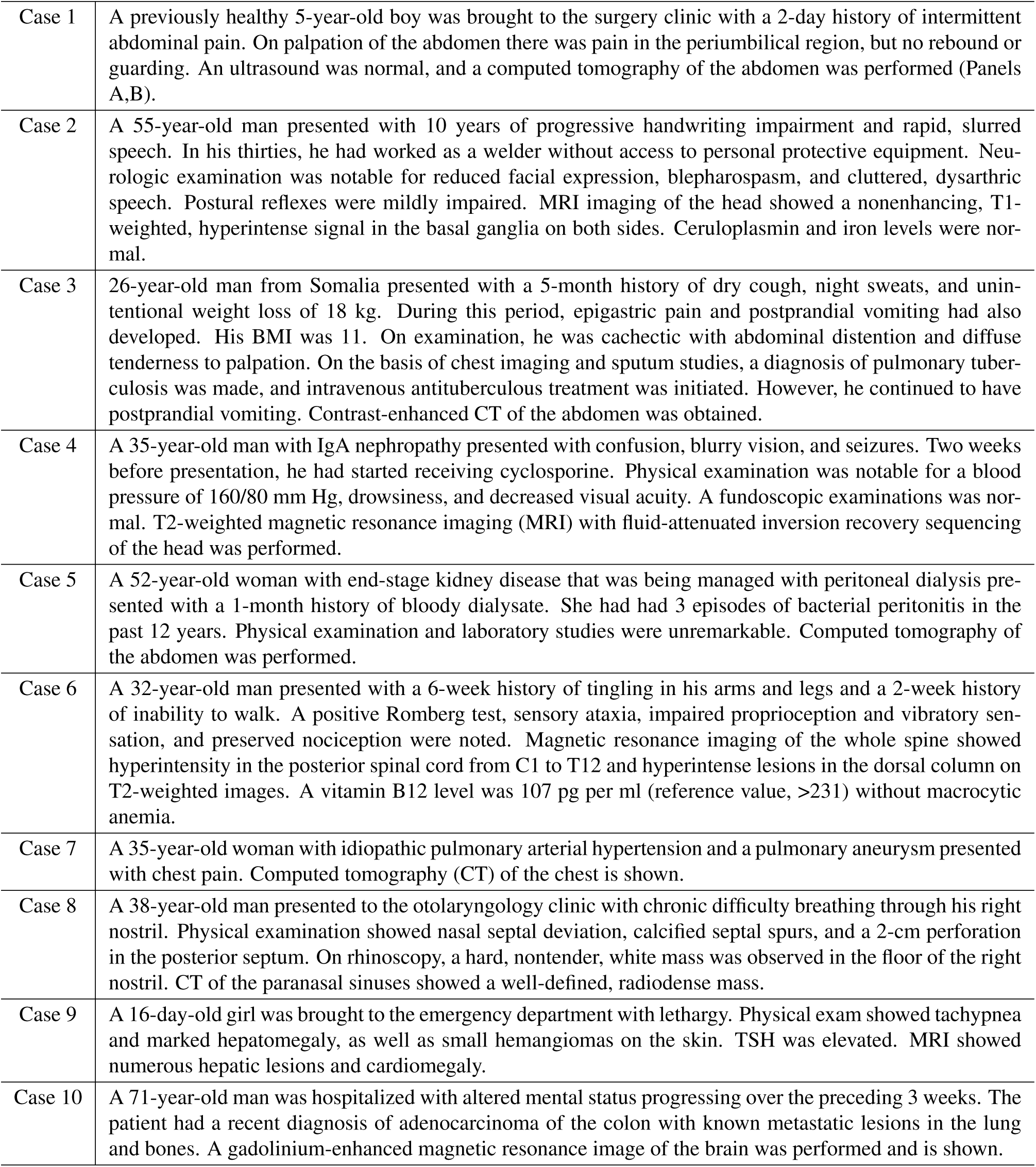
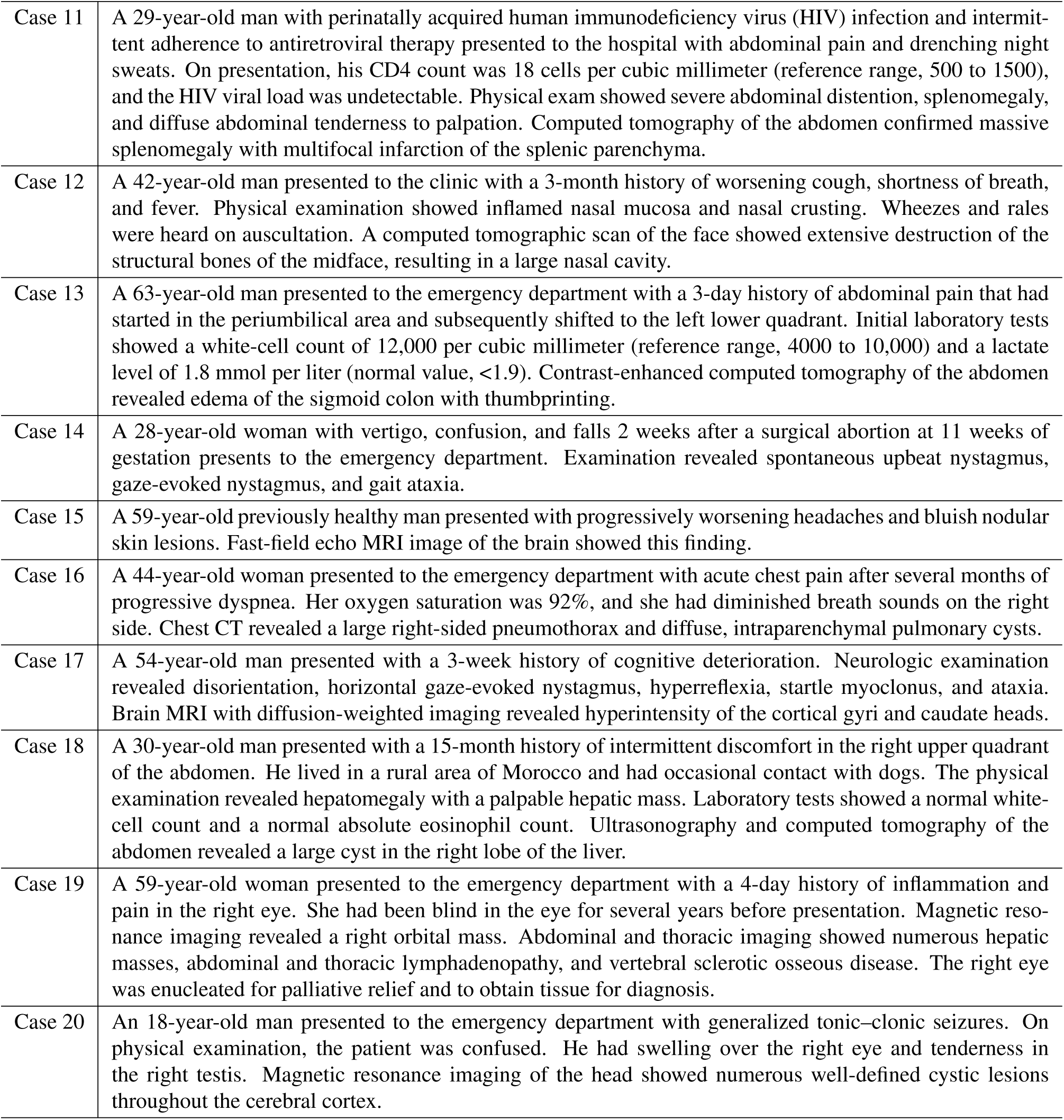
Patient cases included in the study. Description of the 20 diverse patient cases used in the study, which include, for example, pediatric and geriatric patients with various presenting symptoms and diagnostic images.

**Table S4:**
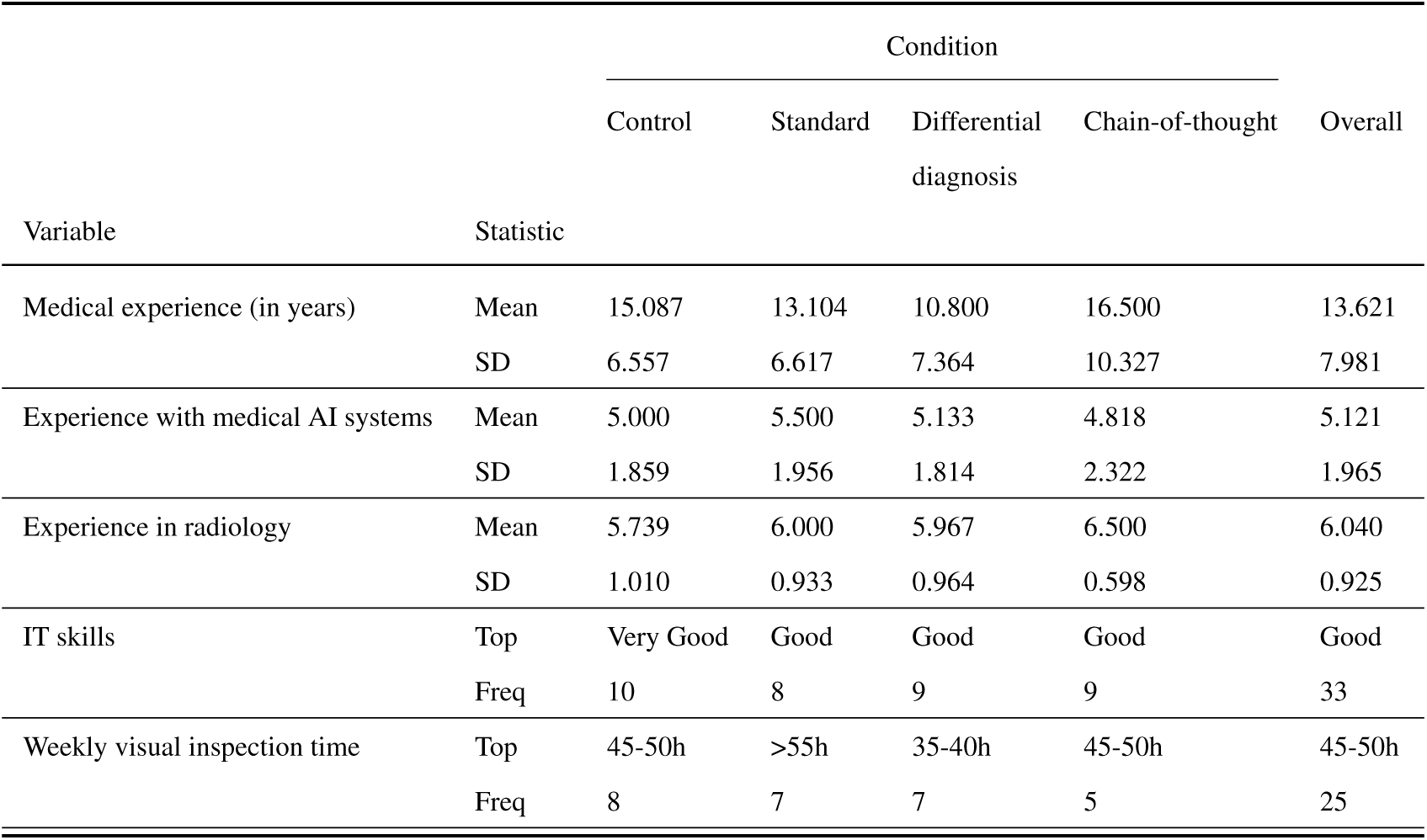
Demographics of study sample by condition. The table presents demographic statistics for study participants across the different experimental conditions. Variables include medical experience (in years), experience with medical AI systems (reported on a 7-point Likert scale), experience in radiology (reported on a 7-point Likert scale), and weekly time spent on visual inspection tasks (reported as discrete time ranges on a 7-item scale). In the table, we report the mean and standard deviation (SD) for continuous variables. For categorical variables, we report the most frequently occurring category (top) and the corresponding frequency (freq). The sample size of the post-study questionnaire varied, as the questionnaire was not fully displayed in two cases where participants selected specific specialties (“neuroradiologist” and “interventional radiology”), leading to *n* = 99 participants with demographic data.

**Table S5:** Number of radiologists by subspecialization. Subspecialization was collected as a multi-option response format, meaning that participants could select multiple subspecializations. As a result, the total sample size in the above table does not equal the *n* = 101 recruited radiologists.

| Subspecialization | Frequency |
| --- | --- |
| General radiology | 57 |
| Abdominal imaging | 24 |
| Interventional radiology | 20 |
| Neuroradiology | 20 |
| Emergency radiology | 19 |
| Musculoskeletal radiology | 18 |
| Mammography / women's imaging | 17 |
| Ultrasound | 15 |
| Nuclear imaging | 11 |
| Head and neck radiology | 9 |
| Cardiothoracic imaging | 8 |
| Pediatric radiology | 5 |
| Other | 4 |

**Table S6:**
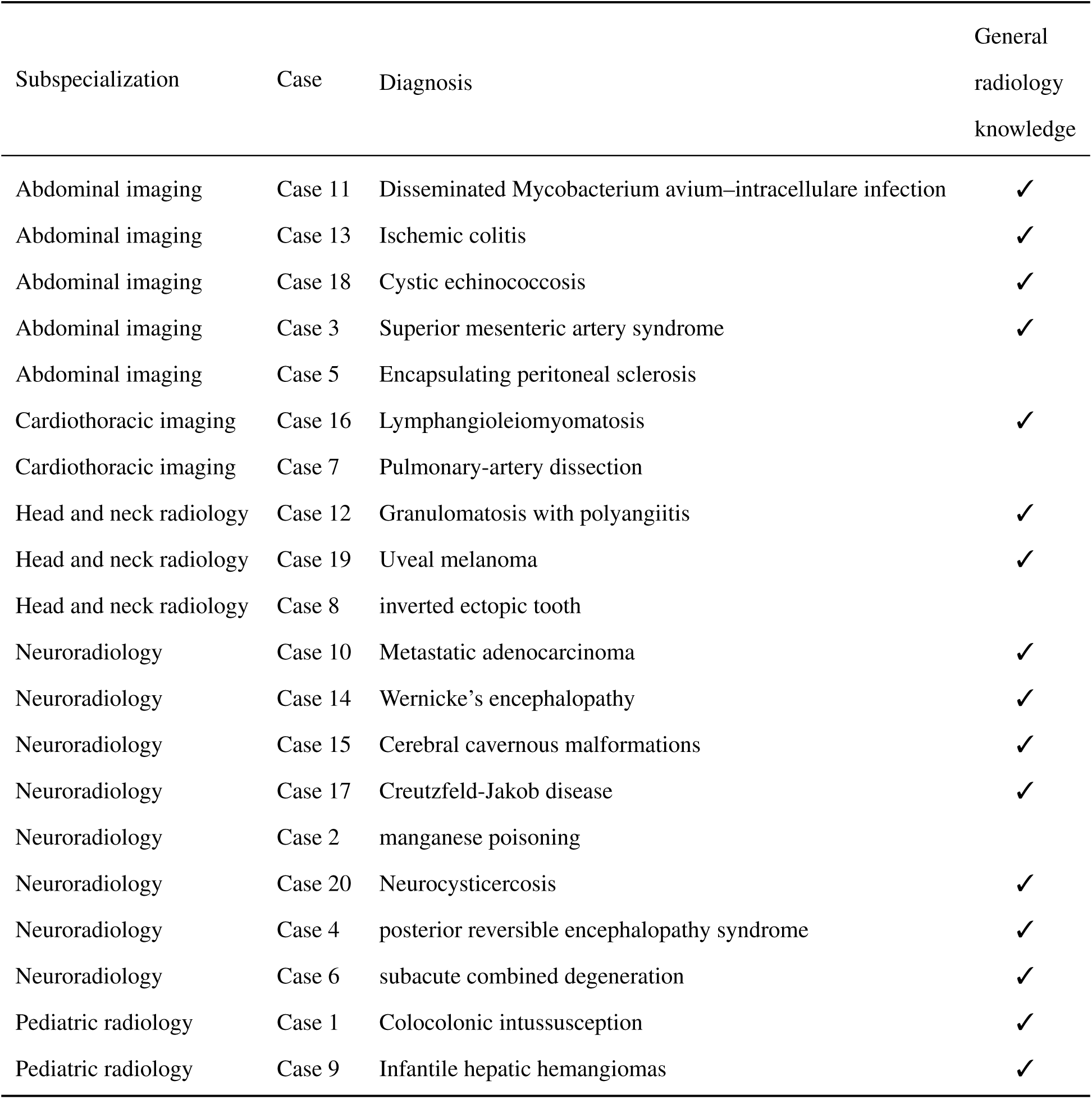
Mapping of patient cases to subspecialization. The specialty name is listed in the first column, followed by the corresponding question and diagnosis. The final column indicates whether the diagnosis is answerable based on general radiology education knowledge, assessed by our panel of radiologists based on common educational resources for radiologist training [25].

**Table S7:** Diagnostic accuracy of the LLM (without human involvement) across different prompting strategies. We estimated a logistic regression at the diagnostic level (i.e., 20 diagnoses *×* 3 conditions). The chain of thought condition is used as the reference category. The odds ratios computed based on the coefficients are 0.750 for the standard condition and 1.000 for the differential diagnosis condition. Hence, considering the intercept, the chain-of-thought approach has a better diagnostic accuracy than the standard prompt and a similar diagnostic accuracy as the differential diagnosis. Abbreviations: s.e., standard error; CI, confidence interval, AIC, Akaike information criterion.

|  | Coef. | s.e. | P-value | 95% CI |
| --- | --- | --- | --- | --- |
| *Intercept* | 1.386 | 0.559 | 0.013 | [0.291; 2.482] |
| *Condition: Standard* | −0.288 | 0.761 | 0.705 | [−1.779; 1.204] |
| *Condition: Differential diagnosis* | 0.000 | 0.791 | 1.000 | [−1.549; 1.549] |
| AIC |  |  |  | 68.526 |
| Obs. (*N*) |  |  |  | 60 |

**Table S8:** Effect of explanation types on diagnostic accuracy. OLS regression explaining the diagnostic accuracy at the physician level by the different conditions. The intercept represents the diagnostic accuracy of humans without LLM support (control condition). The sample size is *n* = 101 (i.e., diagnostic accuracy aggregated at the participant level; *n* = 24 for the control group, *n* = 24 for the standard output group, *n* = 30 for the differential diagnosis group, and *n* = 23 for the chain-of-thought group). Abbreviations: s.e., standard error; CI, confidence interval, AIC, Akaike information criterion.

|  | Coef. | s.e. | *P*-value | 95% CI |
| --- | --- | --- | --- | --- |
| *Intercept* | 0.617 | 0.024 | < 0.001 | [0.568; 0.665] |
| *Condition: Standard* | 0.050 | 0.034 | 0.150 | [−0.018; 0.118] |
| *Condition: Differential diagnosis* | 0.025 | 0.033 | 0.446 | [−0.040; 0.090] |
| *Condition: Chain-of-thought* | 0.122 | 0.035 | 0.001 | [0.053; 0.192] |
| AIC |  |  |  | −138.835 |
| Obs. (*N*) |  |  |  | 101 |

**Table S9:** Benefit of chain-of-thought explanations on diagnostic accuracy. OLS regression of diagnostic accuracy at the physician level on conditions with chain-of-thought as baseline (=intercept). The sample size is *n* = 101 (i.e., diagnostic accuracy aggregated at the participant level; *n* = 24 for the control group, *n* = 24 for the standard output group, *n* = 30 for the differential diagnosis group, and *n* = 23 for the chain-of-thought group). Abbreviations: s.e., standard error; CI, confidence interval, AIC, Akaike information criterion.

|  | Coef. | s.e. | *P*-value | 95% CI |
| --- | --- | --- | --- | --- |
| *Intercept* | 0.739 | 0.025 | < 0.001 | [0.690; 0.789] |
| *Condition: Control* | −0.122 | 0.035 | 0.001 | [−0.192; −0.053] |
| *Condition: Standard* | −0.072 | 0.035 | 0.040 | [−0.142; −0.003] |
| *Condition: Differential diagnosis* | −0.097 | 0.033 | 0.004 | [−0.163; −0.032] |
| AIC |  |  |  | −138.835 |
| Obs. (*N*) |  |  |  | 101 |

**Table S10:** Extended OLS regression of diagnostic accuracy with physician-level controls. Reference category is the control group. In the post-survey questionnaire, the sample size of participants across conditions totals *n* = 99, as the questionnaire was not displayed in two cases where participants selected specific specialties (‘neuroradiologist’ and ‘interventional radiology’). Hence, the two cases were dropped for the analysis. The sample size is *n* = 99 (i.e., diagnostic accuracy aggregated at the participant level; *n* = 23 for the control group, *n* = 24 for the standard output group, *n* = 30 for the differential diagnosis group, and *n* = 22 for the chain-of-thought group). Abbreviations: s.e., standard error; CI, confidence interval, AIC, Akaike information criterion.

|  | Coef. | s.e. | P-value | 95% CI |
| --- | --- | --- | --- | --- |
| *Intercept* | 0.604 | 0.123 | < 0.001 | [0.359; 0.850] |
| *Condition: Standard* | 0.070 | 0.040 | 0.081 | [−0.009; 0.149] |
| *Condition: Differential diagnosis* | 0.039 | 0.039 | 0.325 | [−0.039; 0.117] |
| *Condition: Chain-of-thought* | 0.151 | 0.041 | < 0.001 | [0.070; 0.232] |
| *Medical experience* | 0.001 | 0.002 | 0.707 | [−0.003; 0.004] |
| *Experience in radiology* | −0.001 | 0.001 | 0.301 | [−0.003; 0.001] |
| *Experience with medical AI systems* | −0.000 | 0.000 | 0.519 | [−0.001; 0.001] |
| *Weekly visual inspection time: 30–35h* | 0.077 | 0.108 | 0.478 | [−0.138; 0.292] |
| *Weekly visual inspection time: 35–40h* | 0.043 | 0.101 | 0.667 | [−0.157; 0.244] |
| *Weekly visual inspection time: 40–45h* | 0.067 | 0.100 | 0.508 | [−0.133; 0.266] |
| *Weekly visual inspection time: 45–50h* | 0.076 | 0.097 | 0.441 | [−0.118; 0.269] |
| *Weekly visual inspection time: 50–55h* | 0.039 | 0.100 | 0.698 | [−0.159; 0.237] |
| *Weekly visual inspection time: >55h* | 0.047 | 0.102 | 0.648 | [−0.156; 0.249] |
| *IT skills: Good* | 0.004 | 0.046 | 0.935 | [−0.088; 0.095] |
| *IT skills: Very Good* | 0.068 | 0.048 | 0.156 | [−0.027; 0.163] |
| *IT skills: Advanced* | 0.043 | 0.051 | 0.403 | [−0.058; 0.143] |
| *IT skills: Expert* | 0.023 | 0.072 | 0.748 | [−0.120; 0.167] |
| AIC | −116.208 |  |  |  |
| Obs. (*N*) | 99 |  |  |  |

**Table S11:** Extended OLS regression of diagnostic accuracy with advice-level controls. Here, we control for total time (measured in minutes), advice length (measured in the number of words), and answer length (of the reported diagnosis by the physicians in the free-text form, measured in the number of characters). The reference category is the control group. The sample size is *n* = 101 (i.e., diagnostic accuracy aggregated at the participant level; *n* = 24 for the control group, *n* = 24 for the standard output group, *n* = 30 for the differential diagnosis group, and *n* = 23 for the chain-of-thought group). Abbreviations: s.e., standard error; CI, confidence interval, AIC, Akaike information criterion.

|  | Coef. | s.e. | *P*-value | 95% CI |
| --- | --- | --- | --- | --- |
| *Intercept* | 0.544 | 0.041 | < 0.001 | [0.463; 0.625] |
| *Condition: Standard output* | 0.021 | 0.028 | 0.467 | [−0.035; 0.076] |
| *Condition: Differential diagnosis* | −0.048 | 0.017 | 0.005 | [−0.081; −0.015] |
| *Condition: Chain-of-thought* | 0.046 | 0.018 | 0.012 | [0.010; 0.081] |
| *Total time* | 0.000 | 0.000 | 0.125 | [−0.000; 0.001] |
| *Advice length* | 0.000 | 0.000 | 0.030 | [0.000; 0.001] |
| *Answer length* | 0.003 | 0.002 | 0.050 | [−0.000; 0.006] |
| AIC |  |  |  | −141.197 |
| Obs. (*N*) |  |  |  | 101 |

**Table S12:** Subgroup analysis for participants with basic IT skills. OLS regression explaining the diagnostic accuracy at the physician level by the conditions for the subgroup of participants that have basic IT skills (i.e., lower or equal to “Good”). Reference category is the control group. The sample size is *n* = 43 (i.e., diagnostic accuracy aggregated at the participant level; *n* = 10 for the control group, *n* = 11 for the standard output group, *n* = 10 for the differential diagnosis group, and *n* = 12 for the chain-of-thought group). Abbreviations: s.e., standard error; CI, confidence interval, AIC, Akaike information criterion.

|  | Coef. | s.e. | *P*-value | 95% CI |
| --- | --- | --- | --- | --- |
| *Intercept* | 0.575 | 0.037 | < 0.001 | [0.500; 0.650] |
| *Condition: Standard* | 0.025 | 0.051 | 0.628 | [−0.079; 0.129] |
| *Condition: Differential diagnosis* | 0.090 | 0.052 | 0.094 | [−0.016; 0.196] |
| *Condition: Chain-of-thought* | 0.167 | 0.050 | 0.002 | [0.065; 0.268] |
| AIC |  |  |  | −58.545 |
| Obs. (*N*) |  |  |  | 43 |

**Table S13:** Subgroup analysis for participants with advanced IT skills. OLS regression explaining the diagnostic accuracy at the physician level by the conditions for the subgroup of participants that have advanced IT skills (i.e., better than “Good”). Reference category is the control group. The sample size is *n* = 56 (i.e., diagnostic accuracy aggregated at the participant level; *n* = 24 for the control group, *n* = 24 for the standard output group, *n* = 30 for the differential diagnosis group, and *n* = 23 for the chain-of-thought group). Abbreviations: s.e., standard error; CI, confidence interval, AIC, Akaike information criterion.

|  | Coef. | s.e. | P-value | 95% CI |
| --- | --- | --- | --- | --- |
| *Intercept* | 0.646 | 0.033 | < 0.001 | [0.581; 0.711] |
| *Condition: Standard* | 0.077 | 0.046 | 0.101 | [−0.015; 0.169] |
| *Condition: Differential diagnosis* | −0.016 | 0.042 | 0.701 | [−0.100; 0.068] |
| *Condition: Chain-of-thought* | 0.094 | 0.049 | 0.063 | [−0.005; 0.193] |
| AIC | −77.177 |  |  |  |
| Obs. (*N*) | 56 |  |  |  |

**Table S14:**
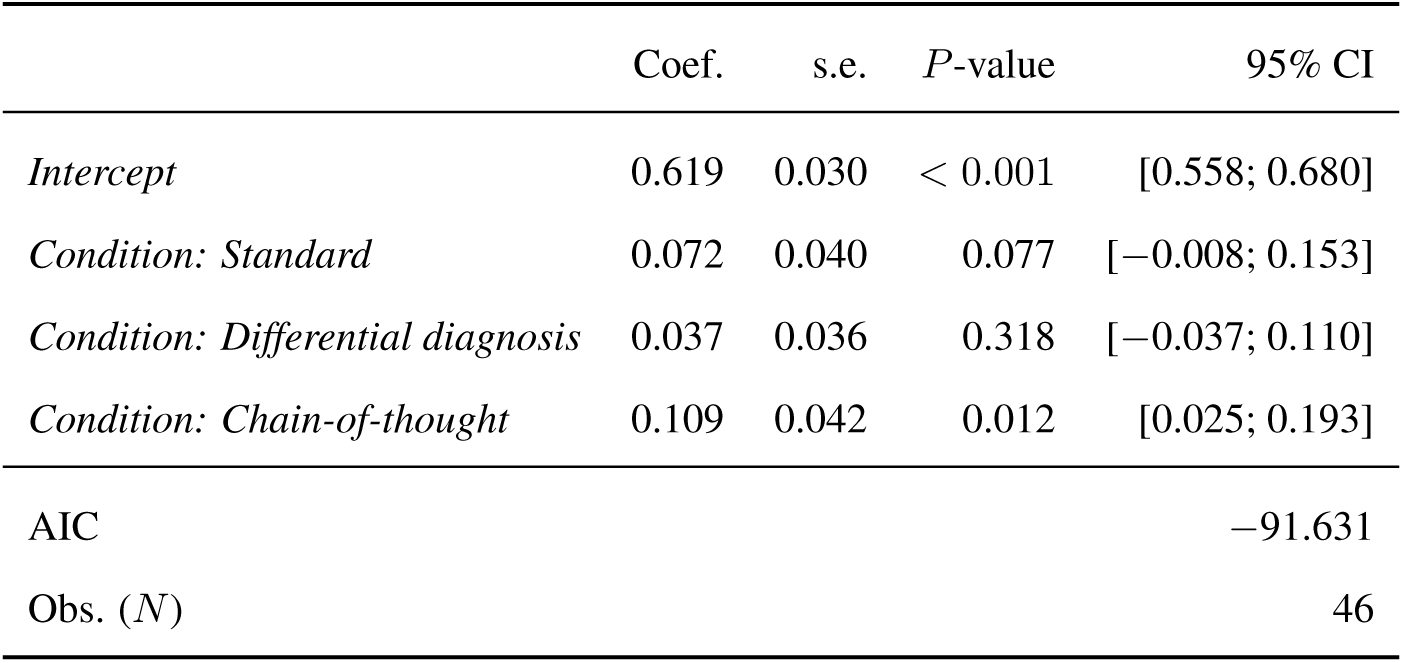
Subgroup analysis for participants with short tenure. OLS regression explaining the diagnostic accuracy at the physician level by the conditions for the subgroup of radiologists with short tenure (less than 12.0 years). Reference category is the control group. The sample size is *n* = 46 (i.e., diagnostic accuracy aggregated at the participant level; *n* = 8 for the control group, *n* = 11 for the standard output group, *n* = 18 for the differential diagnosis group, and *n* = 9 for the chain-of-thought group). Abbreviations: s.e., standard error; CI, confidence interval, AIC, Akaike information criterion.

**Table S15:** Subgroup analysis for participants with long tenure. OLS regression explaining the diagnostic accuracy at the physician level by the conditions for the subgroup of radiologists with long tenure (more than or equal to 12.0 years). Reference category is the control group. The sample size is *n* = 53 (i.e., diagnostic accuracy aggregated at the participant level; *n* = 15 for the control group, *n* = 13 for the standard output group, *n* = 12 for the differential diagnosis group, and *n* = 13 for the chain-of-thought group). Abbreviations: s.e., standard error; CI, confidence interval, AIC, Akaike information criterion.

|  | Coef. | s.e. | *P*-value | 95% CI |
| --- | --- | --- | --- | --- |
| *Intercept* | 0.613 | 0.038 | < 0.001 | [0.537; 0.689] |
| *Condition: Standard* | 0.033 | 0.055 | 0.556 | [−0.078; 0.144] |
| *Condition: Differential diagnosis* | 0.008 | 0.057 | 0.895 | [−0.106; 0.121] |
| *Condition: Chain-of-thought* | 0.137 | 0.055 | 0.017 | [0.025; 0.248] |
| AIC |  |  |  | −49.587 |
| Obs. (*N*) |  |  |  | 53 |

**Table S16:** Subgroup analysis for basic patient cases. OLS regression explaining the diagnostic accuracy at the physician level by the conditions for the subset of basic cases. We split patient cases into basic and complex subsets based on the mean diagnostic accuracy of the control group for each case, where the lower half corresponds to the basic cases. Reference category is the control group. The sample size is *n* = 101 (i.e., diagnostic accuracy aggregated at the participant level; *n* = 24 for the control group, *n* = 24 for the standard output group, *n* = 30 for the differential diagnosis group, and *n* = 23 for the chain-of-thought group). Abbreviations: s.e., standard error; CI, confidence interval, AIC, Akaike information criterion.

|  | Coef. | s.e. | P-value | 95% CI |
| --- | --- | --- | --- | --- |
| *Intercept* | 0.867 | 0.022 | < 0.001 | [0.823; 0.911] |
| *Condition: Standard* | −0.033 | 0.031 | 0.291 | [−0.096; 0.029] |
| *Condition: Differential diagnosis* | −0.057 | 0.030 | 0.060 | [−0.116; 0.002] |
| *Condition: Chain-of-thought* | 0.051 | 0.032 | 0.113 | [−0.012; 0.114] |
| AIC |  |  |  | −157.675 |
| Obs. (*N*) |  |  |  | 101 |

**Table S17:** Subgroup analysis for complex patient cases. OLS regression explaining the diagnostic accuracy at the physician level by the conditions for the subset of complex cases. We split patient cases into basic and complex subsets based on the mean diagnostic accuracy of the control group for each case, where the upper half corresponds to the complex cases. Reference category is the control group. The sample size is *n* = 101 (i.e., diagnostic accuracy aggregated at the participant level; *n* = 24 for the control group, *n* = 24 for the standard output group, *n* = 30 for the differential diagnosis group, and *n* = 23 for the chain-of-thought group). Each diagnostic accuracy is calculated based on the 10 complex patient cases. Abbreviations: s.e., standard error; CI, confidence interval, AIC, Akaike information criterion.

|  | Coef. | s.e. | *P*-value | 95% CI |
| --- | --- | --- | --- | --- |
| *Intercept* | 0.500 | 0.031 | < 0.001 | [0.438; 0.562] |
| *Condition: Standard* | 0.154 | 0.045 | 0.001 | [0.066; 0.243] |
| *Condition: Differential diagnosis* | 0.080 | 0.042 | 0.061 | [−0.004; 0.164] |
| *Condition: Chain-of-thought* | 0.200 | 0.045 | < 0.001 | [0.111; 0.289] |
| AIC |  |  |  | −87.045 |
| Obs. (*N*) |  |  |  | 101 |

**Table S18:** OLS regression controlling for radiologist subspecialization. Here, we follow the OLS regression from the main paper (to estimate the effect compared to the control group) but additionally control for the subspecialization of the physicians (as defined in Supplementary Table S6; subspecializations with no response are dropped from the table). None of the dummies controlling for subspecialization shows significant differences, implying that the differences in radiologists’ backgrounds cannot explain away our main findings. Abbreviations: s.e., standard error; CI, confidence interval, AIC, Akaike information criterion.

|  | Coef. | s.e. | P-value | 95 % CI |
| --- | --- | --- | --- | --- |
| *Intercept* | 0.629 | 0.034 | < 0.001 | [0.562; 0.697] |
| *Condition: Standard output* | 0.048 | 0.040 | 0.241 | [−0.032; 0.128] |
| *Condition: Differential diagnosis* | 0.032 | 0.038 | 0.398 | [−0.043; 0.108] |
| *Condition: Chain-of-thought* | 0.133 | 0.039 | 0.001 | [0.056; 0.211] |
| *Abdominal imaging* | −0.015 | 0.039 | 0.699 | [−0.092; 0.062] |
| *Cardiothoracic imaging* | 0.038 | 0.058 | 0.516 | [−0.077; 0.152] |
| *Emergency radiology* | 0.004 | 0.037 | 0.917 | [−0.070; 0.078] |
| *General radiology* | −0.004 | 0.027 | 0.891 | [−0.058; 0.051] |
| *Head and neck radiology* | −0.022 | 0.063 | 0.733 | [−0.147; 0.104] |
| *Interventional radiology* | −0.048 | 0.035 | 0.175 | [−0.117; 0.022] |
| *Mammography / women's imaging* | −0.005 | 0.037 | 0.883 | [−0.079; 0.068] |
| *Musculoskeletal radiology* | −0.044 | 0.037 | 0.236 | [−0.118; 0.030] |
| *Neuroradiology* | 0.020 | 0.043 | 0.636 | [−0.065; 0.105] |
| *Nuclear Imaging* | −0.012 | 0.045 | 0.791 | [−0.102; 0.078] |
| *Pediatric Radiology* | −0.072 | 0.061 | 0.243 | [−0.194; 0.050] |
| *Ultrasound* | 0.062 | 0.050 | 0.216 | [−0.037; 0.161] |
| *Other* | −0.065 | 0.068 | 0.341 | [−0.200; 0.070] |
| AIC | −120.762 |  |  |  |
| Obs. (*N*) | 101 |  |  |  |

**Table S19:**
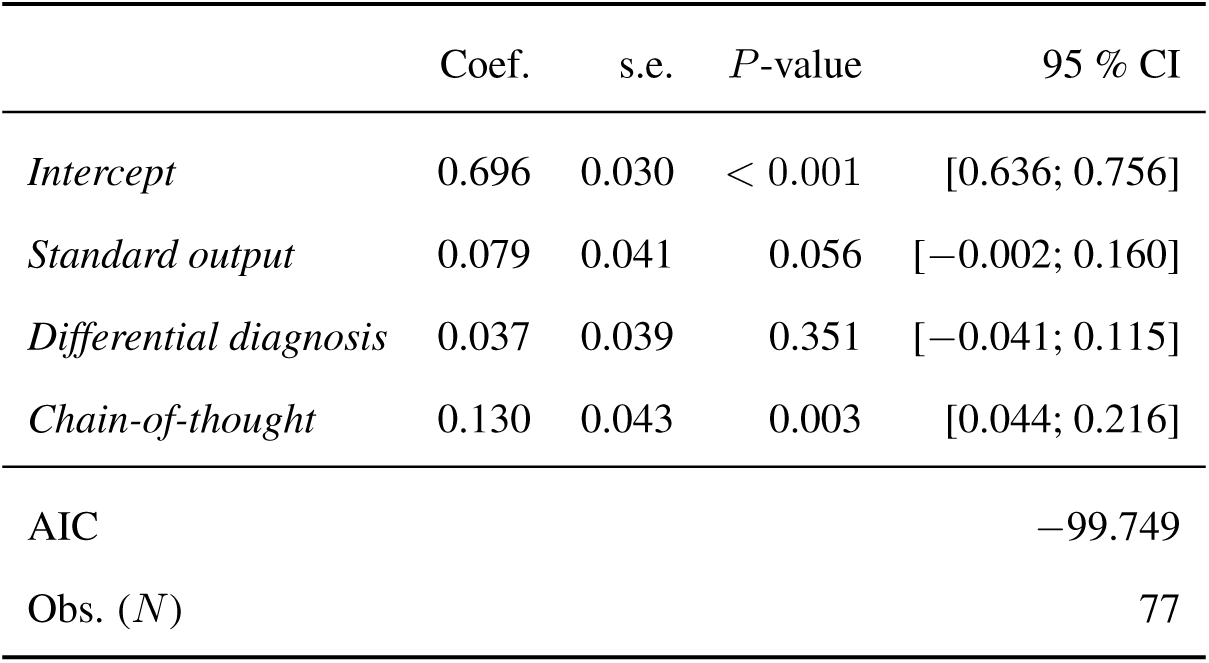
OLS regression explaining the explanation format effect on the diagnostic accuracy for patient cases within participants’ subspecialties. The analysis aims to analyze the scenario in which physicians only diagnose patients in their subspecialty (as defined in Supplementary Table S6). For this, we run the regression (with the control condition as reference) on the relevant, subspecialty-matched subset of the data, which we obtained as follows. The 20 patient cases were assigned to the subspecialties as follows (see Supplementary Table S6): 8 cases to neurology, 3 to head and neck radiology, 5 to abdominal imaging, 2 to cardiothoracic imaging, and 2 to pediatric radiology. Additionally, of the 20 cases, 16 are identified to be answerable based on general radiology knowledge obtained through widespread radiology education resources, such as *“Core Radiology: A Visual Approach to Diagnostic Imaging”* [25], and therefore included in the diagnostic accuracy assessment of all specialized participants. Participants without a focus on general radiology or at least one specialty covered by our patient cases are excluded in this analysis, resulting in a sample size of *n* = 77. We then assess the diagnostic accuracy of the participants based on cases within each participant’s subspecialty. Abbreviations: s.e., standard error; CI, confidence interval; AIC, Akaike information criterion.

|  | Coef. | s.e. | P-value | 95 % CI |
| --- | --- | --- | --- | --- |
| *Intercept* | 0.696 | 0.030 | < 0.001 | [0.636; 0.756] |
| *Standard output* | 0.079 | 0.041 | 0.056 | [−0.002; 0.160] |
| *Differential diagnosis* | 0.037 | 0.039 | 0.351 | [−0.041; 0.115] |
| *Chain-of-thought* | 0.130 | 0.043 | 0.003 | [0.044; 0.216] |
| AIC |  |  |  | −99.749 |
| Obs. (*N*) |  |  |  | 77 |

**Table S20:** OLS regression of explanation format effect on the diagnostic accuracy among general radiologists. The model is estimated on *n* = 57 participants who self-identify as general radiologists. Abbreviations: s.e., standard error; CI, confidence interval; AIC, Akaike information criterion.

|  | Coef. | s.e. | P-value | 95 % CI |
| --- | --- | --- | --- | --- |
| *Intercept* | 0.650 | 0.044 | < 0.001 | [0.562; 0.738] |
| *Condition: Standard output* | 0.009 | 0.054 | 0.862 | [−0.098; 0.117] |
| *Condition: Differential diagnosis* | −0.000 | 0.052 | 1.000 | [−0.104; 0.104] |
| *Condition: Chain-of-thought* | 0.077 | 0.056 | 0.172 | [−0.034; 0.188] |
| AIC |  |  |  | −72.764 |
| Obs. (*N*) |  |  |  | 57 |

**Table S21:** Robustness checks controlling for outliers. OLS regression explaining the winsorized diagnostic accuracy at the physician level by the different conditions. To improve the statistical reliability and reduce the potential bias from outliers or extreme values, we applied winsorization to the diagnostic accuracy at the participant level. We do this by replacing values outside the 5th and 95th percentiles with values at these percentiles, thus limiting the influence of extreme values. Reference category is the control group. The sample size is *n* = 101 (i.e., diagnostic accuracy aggregated at the participant level; *n* = 24 for the control group, *n* = 24 for the standard output group, *n* = 30 for the differential diagnosis group, and *n* = 23 for the chain-of-thought group). Abbreviations: s.e., standard error; CI, confidence interval, AIC, Akaike information criterion.

|  | Coef. | s.e. | *P*-value | 95% CI |
| --- | --- | --- | --- | --- |
| *Intercept* | 0.625 | 0.021 | < 0.001 | [0.584; 0.666] |
| *Condition: Standard* | 0.052 | 0.029 | 0.079 | [−0.006; 0.110] |
| *Condition: Differential diagnosis* | 0.015 | 0.028 | 0.591 | [−0.040; 0.070] |
| *Condition: Chain-of-thought* | 0.114 | 0.030 | < 0.001 | [0.055; 0.173] |
| AIC |  |  |  | −171.332 |
| Obs. (*N*) |  |  |  | 101 |

**Table S22:** Mixed-effects model. We estimated a mixed-effects model to control for heterogeneity across physicians and cases. Reference category is the control group. As we control for variables assessed in the questionnaire, which have not been displayed in two cases, the sample size is reduced to *N* = 1980 = 2020 *−* 20 *×* 2 assessments (*n* = 460 for the control group, *n* = 480 for the standard output group, *n* = 600 for the differential diagnosis group, and *n* = 440 for the chain-of-thought group), which 20 observations belonging to each participant. Abbreviations: s.e., standard error; CI, confidence interval, AIC, Akaike information criterion.

|  | Coef. | s.e. | P-value | 95% CI |
| --- | --- | --- | --- | --- |
| *Intercept* | 1.036 | 0.823 | 0.208 | [−0.577; 2.648] |
| *Condition: Standard* | 0.548 | 0.248 | 0.027 | [0.062; 1.034] |
| *Condition: Differential diagnosis* | 0.095 | 0.242 | 0.695 | [−0.379; 0.569] |
| *Condition: Chain-of-thought* | 1.141 | 0.259 | < 0.001 | [0.633; 1.650] |
| *Medical experience* | 0.001 | 0.011 | 0.922 | [−0.021; 0.023] |
| *Experience in radiology* | −0.008 | 0.007 | 0.256 | [−0.021; 0.006] |
| *Experience with medical AI systems* | −0.003 | 0.003 | 0.316 | [−0.009; 0.003] |
| *Weekly visual inspection time: 30–35h* | 0.738 | 0.663 | 0.265 | [−0.561; 2.038] |
| *Weekly visual inspection time: 35–40h* | 0.361 | 0.610 | 0.554 | [−0.834; 1.556] |
| *Weekly visual inspection time: 40–45h* | 0.551 | 0.607 | 0.364 | [−0.639; 1.741] |
| *Weekly visual inspection time: 45–50h* | 0.535 | 0.590 | 0.364 | [−0.621; 1.691] |
| *Weekly visual inspection time: 50–55h* | 0.299 | 0.604 | 0.621 | [−0.884; 1.482] |
| *Weekly visual inspection time: >55h* | 0.314 | 0.617 | 0.611 | [−0.896; 1.524] |
| *IT skills: Good* | 0.070 | 0.285 | 0.805 | [−0.488; 0.628] |
| *IT skills: Very Good* | 0.555 | 0.299 | 0.063 | [−0.030; 1.140] |
| *IT skills: Advanced* | 0.516 | 0.316 | 0.103 | [−0.104; 1.136] |
| *IT skills: Expert* | 0.330 | 0.452 | 0.465 | [−0.556; 1.215] |
| AIC | 1780.944 |  |  |  |
| Obs. (*N*) | 1980 |  |  |  |
| Variance explained by participant IDs | 0.235 |  |  |  |
| Variance explained by case IDs | 2.275 |  |  |  |

**Table S23:** Quasi-binomial regression. We estimated quasi-binomial regression, which may limit the interpretability of the estimated effect sizes but which can better accommodate for the fact that the diagnostic accuracy is in the range between 0% and 100%. Here, we fitted a generalized linear model with a logistic link function, using a binomial error distribution to model the diagnostic accuracy of responses based on the conditions from our experiment. We further adjusted for overdispersion by scaling the standard errors using a dispersion factor estimated from the Pearson *χ*^2^-statistic. Reference category is the control group. The sample size is *n* = 101 (i.e., diagnostic accuracy aggregated at the participant level; *n* = 24 for the control group, *n* = 24 for the standard output group, *n* = 30 for the differential diagnosis group, and *n* = 23 for the chain-of-thought group). Abbreviations: s.e., standard error; CI, confidence interval, AIC, Akaike information criterion.

|  | Coef. | s.e. | *P*-value | 95% CI |
| --- | --- | --- | --- | --- |
| *Intercept* | 0.475 | 0.105 | < 0.001 | [0.269; 0.681] |
| *Condition: Standard output* | 0.218 | 0.151 | 0.149 | [−0.078; 0.514] |
| *Condition: Differential diagnosis* | 0.107 | 0.142 | 0.450 | [−0.171; 0.385] |
| *Condition: Chain-of-thought* | 0.566 | 0.159 | < 0.001 | [0.255; 0.877] |
| AIC | 95.364 |  |  |  |
| Obs. (*N*) | 101 |  |  |  |

**Table S24:** Effect on decision time. OLS regression explaining the average time to answer each case across the different conditions, leading to *N* = 80 observations (= 20 cases *×* 4 conditions). Reference category is the control group. Abbreviations: s.e., standard error; CI, confidence interval, AIC, Akaike information criterion.

|  | Coef. | s.e. | *P*-value | 95% CI |
| --- | --- | --- | --- | --- |
| *Intercept* | 79.369 | 30.649 | 0.012 | [18.327; 140.411] |
| *Condition: Standard* | 13.117 | 43.344 | 0.763 | [−73.209; 99.443] |
| *Condition: Differential diagnosis* | 68.332 | 43.344 | 0.119 | [−17.994; 154.659] |
| *Condition: Chain-of-thought* | 57.825 | 43.344 | 0.186 | [−28.501; 144.152] |
| AIC | 1018.199 |  |  |  |
| Obs. (*N*) | 80 |  |  |  |

**Table S25:** Effect of decision time (compared to the standard group). OLS regression explaining the average time to answer each case across the different conditions. The standard output group is set as the reference group, leading to *N* = 60 observations (= 20 cases *×*3 treatment groups). Abbreviations: s.e., standard error; CI, confidence interval, AIC, Akaike information criterion.

|  | Coef. | s.e. | *P*-value | 95 % CI |
| --- | --- | --- | --- | --- |
| *Intercept* | 92.485 | 34.900 | 0.010 | [22.599; 162.372] |
| *Condition: Differential diagnosis* | 55.216 | 49.356 | 0.268 | [−43.619; 154.050] |
| *Condition: Chain-of-thought* | 44.709 | 49.356 | 0.369 | [−54.126; 143.543] |
| AIC | 779.238 |  |  |  |
| Obs. (*N*) | 60 |  |  |  |

**Table S26:** Logistic regression at the assessment level. Logistic regression at the assessment level (=1 if the correct diagnosis was given by a physician for a specific case, 0 otherwise) explaining the diagnostic accuracy by the different conditions. Reference category is the control group. The sample size consists of *N* = 2020 observations, each participant measured 20 times (*N* = 480 for the control group, *N* = 480 for the standard output group, *N* = 600 for the differential diagnosis group, and *N* = 460 for the chain-of-thought group). Abbreviations: s.e., standard error; CI, confidence interval, AIC, Akaike information criterion.

|  | Coef. | s.e. | *P*-value | 95% CI |
| --- | --- | --- | --- | --- |
| *Intercept* | 0.769 | 0.098 | < 0.001 | [0.577; 0.961] |
| *Condition: Standard* | 0.296 | 0.143 | 0.039 | [0.015; 0.577] |
| *Condition: Differential diagnosis* | 0.054 | 0.132 | 0.680 | [−0.205; 0.314] |
| *Condition: Chain-of-thought* | 0.672 | 0.154 | < 0.001 | [0.371; 0.974] |
| AIC | 2340.802 |  |  |  |
| Obs. (*N*) | 2020 |  |  |  |

**Table S27:** Logistic regression controlling for correct explanations and correct diagnosis in the LLM advice. Logistic regression at the assessment level (=1 if the correct diagnosis was given by a physician for a specific case, 0 otherwise) explaining the diagnostic accuracy by the different conditions. Here, we separately control for (a) whether the suggested diagnosis of the LLM is correct or (b) whether the explanation generated by the LLM is correct. The diagnosis in the differential diagnosis group is considered correct if the correct diagnosis is one of the five presented options. The chain-of-thought group served as the reference group. We omit the control group because the control did not receive LLM advice, making it impossible to control for the correctness of that advice. The sample size consists of *N* = 1540 observations, for which 20 observations belong to each participant (*N* = 480 for the standard output group, *N* = 600 for the differential diagnosis group, and *N* = 460 for the chain-of-thought group). Abbreviations: s.e., standard error; CI, confidence interval, AIC, Akaike information criterion.

|  | Coef. | s.e. | P-value | 95% CI |
| --- | --- | --- | --- | --- |
| *Intercept* | −0.619 | 0.171 | < 0.001 | [−0.954; −0.285] |
| *Condition: Standard* | −0.504 | 0.209 | 0.016 | [−0.914; −0.094] |
| *Condition: Differential diagnosis* | −1.014 | 0.188 | < 0.001 | [−1.383; −0.646] |
| *Correct explanation (True = 1): 1* | 0.462 | 0.162 | 0.004 | [0.144; 0.779] |
| *Correct diagnosis (True = 1): 1* | 2.900 | 0.168 | < 0.001 | [2.571; 3.229] |
| AIC | 1263.086 |  |  |  |
| Obs. (*N*) | 1540 |  |  |  |

**Table S28:** Regression controlling for the characteristics of radiological images. Logistic regression at the assessment level (=1 if the correct diagnosis was given by a physician for a specific case, 0 otherwise) explaining the diagnostic accuracy by the different conditions and image characteristics. Here, we include (a) the number of images shown for a patient case, (b) the source of the image(s) (i.e., MRI or CT [CT serves as a baseline in the regression]), and (c) the body region shown in the image(s) (abdomen serves as a baseline for the regression here). By exponentiating the coefficients, the odds ratios of 1.60 for the standard condition, 1.24 for the differential condition, and 2.37 for the chain-of-thought condition are obtained. The sample size consists of *N* = 2020 observations, with 20 observations per participant (*N* = 480 for the control group, *N* = 480 for the standard group, *n* = 600 for the differential diagnosis group, and *n* = 460 for the chain-of-thought group). Abbreviations: s.e., standard error; CI, confidence interval.

|  | Coef. | s.e. | P-value | 95 % CI |
| --- | --- | --- | --- | --- |
| *Condition: Standard* | 0.470 | 0.137 | 0.001 | [0.201; 0.739] |
| *Condition: Differential diagnosis* | 0.216 | 0.125 | 0.085 | [−0.030; 0.461] |
| *Condition: Chain-of-thought* | 0.862 | 0.148 | < 0.001 | [0.572; 1.152] |
| *Image: Count* | 0.045 | 0.075 | 0.546 | [−0.102; 0.192] |
| *Source: MRI* | 0.729 | 0.141 | < 0.001 | [0.451; 1.006] |
| *Body region: Head* | 0.192 | 0.136 | 0.159 | [−0.075; 0.460] |
| *Body region: Spine* | 1.806 | 0.488 | < 0.001 | [0.849; 2.762] |
| *Body region: Thorax* | 0.719 | 0.176 | < 0.001 | [0.375; 1.064] |
| AIC |  |  |  | 2267.722 |
| Obs. (*N*) |  |  |  | 2020 |

### Supplementary Materials

#### Impact of explanation formats on decision time

Physicians randomized to LLM support spent more time per case, but in the order of around 1–2 minutes (see Supplementary Figure S6). The control group had the lowest decision time with 79.4 seconds per patient case. To calculate the effect on decision time for the different explanation formats compared to the control group, we again estimated an OLS regression model (Supplementary Table S24). The decision time per patient case was 92.5 seconds for the standard output, similar to the control group (difference = 13.1 seconds, 95% CI = [*−*73.2, 99.4], *P* = 0.763). The differential diagnosis group and the chain-of-thought group had an average decision time per patient case of 147.6 and 137.2 seconds (difference = 68.3 seconds 95% CI = [*−*18.0, 154.7], *P* = 0.119; difference = 57.8 seconds, 95% CI = [*−*28.5, 144.2], *P* = 0.186; respectively). Additionally, we estimated an OLS regression model to compare the standard output group against (i) the differential diagnosis group and (ii) the chain-of-thought explanation group (Supplementary Table S25). The differential diagnosis group and the chain-of-thought group had a similar average decision time per patient case compared to the standard output group (difference = 61.8 seconds, 95% CI = [*−*12.5, 136.1], *P* = 0.102; and difference = 51.3 seconds, 95% CI = [*−*23.0, 125.6], *P* = 0.173; respectively).

We now turn to the economic implications of our findings. While decision time was recorded in our study, it was not the primary focus; instead, the emphasis lies on how explanation formats influence diagnostic performance. Importantly, our main results remain robust even when controlling for decision time, reinforcing the benefit of chain-of-thought explanations. Although we report time-related outcomes, we are cautious not to overinterpret them in economic terms. In clinical practice, the costs associated with misdiagnoses—such as delayed or inappropriate treatment—can far exceed the marginal costs of LLM support, often by several orders of magnitude.

Thus, we highlight the large improvements in diagnostic accuracy from optimizing the explanation format of medical LLMs.

To see this, consider the following back-of-the-envelope analysis, where all assumptions are deliberately conservative. We begin by assuming a scenario of 100 daily diagnostic decisions per physician. With a documented improvement in diagnostic accuracy of 12.2 percentage points, this corresponds to an expected reduction of misdiagnoses by 100 *×* 0.122 = 12.2 cases. To quantify how each avoided misdiagnosis averts downstream costs, we consider two scenarios. First, we use a highly conservative estimate. If each misdiagnosis costs $100, then the avoided costs amount to 12 *×* 100 = 1,200 USD. The cost of processing explanations is given by 100 *×* 2 = 200 minutes (assuming an excess time 2 min/diagnosis), or 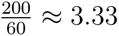 hours. At a labor cost of $200 per hour, this results in 3.33 *×* 200 = 666.67 USD in added labor. The net financial benefit is therefore 1,200 *−* 666.67 = 533.33 USD per physician per day. Second, we assume make the conservative assumption that each misdiagnosis costs $1,000. Then, the avoided costs are 12 *×* 1,000 = 12,000 USD. Subtracting the same explanation processing cost of 666.67 USD yields a net benefit of 12,000 *−* 666.67 = 11,333.33 USD per physician per day.

Together, the above results demonstrate that, even under conservative assumptions, incorporating chain-of-thought explanations into diagnostic workflows is linked to a net gain. The analysis underscores that the downstream consequences of misdiagnosis substantially outweigh the modest time investment required to interpret model-generated explanations, indicating a favorable tradeoff in terms of cost effectiveness.

#### Estimating the maximum expected performance as an upper bound of the achievable accuracy in each condition

To assess the upper bound of participant accuracy under different explanation strategies, we define and calculate the *maximum expected performance* (MEP). The MEP quantifies the highest possible accuracy achievable by participants under two assumptions of how LLM advice is translated into decisions: (1) when the LLM provides a correct diagnosis and the participant adheres to it, the participant is also correct; (2) when the LLM provides an incorrect diagnosis but the participant overrules it, we assume the participant responds correctly. Formally, we thus compute MEP via

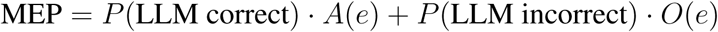

where *A*(*e*) denotes the adherence rate for explanation type *e* given a correct LLM answer, and *O*(*e*) = 1 *− A*(*e*) denotes the overruling rate given an incorrect LLM answer. The values of *P* (LLM correct) and *P* (LLM incorrect) are set based on the observed accuracy of the LLM at 80%.

The empirical rates for adherence were obtained from Fig. 3. Specifically, when the LLM’s diagnosis was incorrect, adherence rates were 30% for the chain-of-thought group, 76% for the differential diagnosis group, and 28% for the standard group. When the diagnosis was correct, adherence rates were 92%, 83%, and 89%, respectively. Substituting these values, we obtain:

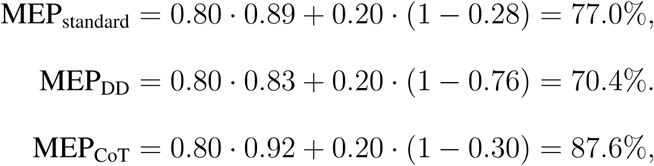

These estimates thus represent upper bounds on participant performance, suggesting that, due to differences in adherence, the potential gain from the CoT condition is larger than for the differential diagnosis condition. Importantly, the observed accuracies in the experiments fall within these upper bounds, thereby reinforcing the internal consistency of our analyses.

## Footnotes

1 We follow the terminology of literature on human-computer interaction and use the term “override” to describe the scenario in which a human is not adhering to an AI-generated recommendation, regardless of whether this behavior is correct [29].

